# Refining uncertainty about the TAK-003 dengue vaccine with a multi-level model of clinical efficacy trial data

**DOI:** 10.1101/2025.07.31.25332513

**Authors:** Manar Alkuzweny, Guido España, T. Alex Perkins

## Abstract

**Background:** A safe and effective vaccine that can be universally administered would represent a major advance in efforts to control dengue. Takeda’s TAK-003 is a potential candidate for such a vaccine. However, phase-III trial results suggest that TAK-003 confers differential protection by outcome, serotype, and serostatus. Published analyses estimated these stratified vaccine efficacies independently, ignoring the fact that some aspects of the disease process are shared across particular stratifications.

**Methods:** We addressed these shortcomings by building a multi-level model that pooled all publicly available trial data to estimate parameters that characterize this vaccine’s profile.

**Results:** We found that protection varied by both serotype and serostatus, with initial protection against symptomatic disease ranging from a median of 99.6% (95% credible interval [CrI]: 96.5, 100.0) among seronegatives infected with DENV-2 to 26.7% (95% CrI: −8.2, 54.9) among seronegatives infected with DENV-3. We found that initial protection against hospitalized disease among those with symptomatic disease ranged from 98.7% (95% CrI: 90.1, 100.0) among seropositives infected with DENV-2 to −72.7% (95% CrI: −149.4, −4.0) among seronegatives infected with DENV-3. Importantly, our estimate of the latter is less uncertain than a previously published efficacy estimate (−87.9%, 95% CI: −573.4, 47.6).

**Conclusions:** Our results demonstrate that inferences based on complex data from a clinical trial can be sensitive to model structure. Additionally, they reinforce the recommendation from the World Health Organization that use of this vaccine is best suited to high-transmission settings.

## INTRODUCTION

Dengue is a viral disease transmitted by *Aedes* mosquitoes. Over half of the world’s population lives in areas at risk of dengue virus (DENV) transmission, and nearly 100 million apparent infections occur annually (1,2). There are four antigenically distinct serotypes of dengue virus that can result in individuals becoming infected up to four times, with the second infection posing the greatest risk of severe outcomes (3). Due to this complex natural history, it has proven challenging to develop a vaccine that confers adequate protection against all four serotypes without increasing the risk of severe disease among seronegative individuals (4–6). Notably, the first licensed dengue vaccine, Sanofi Pasteur’s Dengvaxia, increases the risk of severe disease in vaccinated seronegatives, which has led to low uptake globally (7–13). As such, there remains an unmet need for a safe and effective dengue vaccine that can be universally administered.

One candidate that could potentially fill this gap is TAK-003, a live-attenuated tetravalent vaccine developed by Takeda that utilizes DENV-2 for its genetic backbone (14). This vaccine has been recommended for use in high-transmission settings by the World Health Organization (15) and is now being administered in several countries (16–25). Trial results suggest that the vaccine offers differential protection depending on the infecting serotype and an individual’s baseline serostatus. In particular, published vaccine efficacy estimates indicate higher protection against DENV-2, and potentially enhanced risk of disease for vaccinated seronegatives infected with DENV-3 or DENV-4 (16–19,25,26). However, these estimates come with broad uncertainty, in part due to relatively low numbers of DENV-3 and DENV-4 cases observed during the trial (16–19,12,25). Given that TAK-003 is now being rolled out for public health use, there is an urgent need to resolve outstanding uncertainties about this vaccine’s characteristics (27). While this has been partially addressed by work that aimed to estimate vaccine efficacy at finer stratifications than previously published (28), an analysis that aims to refine uncertainty by simultaneously utilizing all available data has not yet been performed.

One way to address the broad uncertainty of previous estimates is to jointly analyze trial data with a multi-level model. This approach can lead to more precise estimates by specifying aspects of the data that may be correlated due to similarities in their underlying causal processes. For example, individuals in the same country should be subject to similar factors that affect whether their disease is reported, and individuals with the same serostatus should share a similar probability of disease given infection (29). Assuming that individuals who share baseline characteristics (e.g., country or serostatus) experience independent probabilities of becoming infected or hospitalized is not realistic, yet those assumptions were implicitly made in the original analysis of trial data (16–19,25). In addition, although limited information about the stratification of trial data has been made publicly available (16–19,25), any aggregation of data could increase the risk of bias and inflate uncertainty if the model fails to account for heterogeneities that shape the data (30). For example, serotype-specific forces of infection and criteria for hospitalization are both known to vary tremendously across settings (31,32). While previous work has addressed the impact of accounting for age on serotype- and serostatus-specific estimates, accounting for such setting-specific differences to resolve reported uncertainty has yet to be done. If we disregard these differences, we risk mistaking the effects of these heterogeneities as inflated uncertainty about the vaccine. A multi-level model allows us to preserve stratifications in the data and specify some parameters as uniform across stratifications (e.g., vaccine characteristics) and other parameters as stratum-specific (e.g., country-specific force of infection) (33).

In this study, we jointly leveraged all publicly available clinical trial data at their most granular resolution to obtain refined estimates of the TAK-003 vaccine’s profile. Specifically, the data that we used were stratified by clinical endpoint, age, serostatus, country, serotype, and time interval, although not by all at once (16–19,25). For example, country-level serotype-specific data were not stratified by baseline serostatus or age, and they were only available for one time interval. By specifying the model’s structure in a way that is consistent with these aspects of how the data were generated and assuming a leaky vaccine (34), we were able to obtain estimates of per-exposure protection. This differs from efficacy in that it is an individual-level parameter that can be used to predict vaccine impact in contexts outside of the trial (35). We report our parameter estimates, assess their agreement with trial data, and comment on their implications.

## METHODS

### Data description

Data on cases and hospitalizations among eligible participants were made available for varying time intervals between 4 and 57 months after the first of two doses of the vaccine, which were administered three months apart (16). Participants were considered eligible if they were 4–16 years old at the start of the trial and in good health (16). Cases were defined as clinically suspected dengue (body temperature >38 °C for two out of three consecutive days) confirmed via positive serotype-specific reverse-transcriptase polymerase chain reaction (RT-PCR) (16). Hospitalizations were recorded from hospitals included as study sites in the trial and were a subset of virologically confirmed dengue cases. Additionally, data were published on the proportion of participants who were seropositive at baseline by country (16).

Importantly for our analysis, data were available at differing levels of aggregation by outcome (outcome type and infecting serotype) and group (baseline serostatus, age, and country) (Table 1). Additionally, each of these pieces of data were available at different time intervals. For example, serotype-specific case data stratified by serostatus and treatment arm were available in five intervals from 4 to 57 months after the first dose, while case data stratified by country and arm were available for only a single time interval from 1 to 39 months after the first dose (Table 1). To accommodate all the varied aggregation levels at which data were available, we developed an approach that was flexible enough to aggregate granular model predictions to match more aggregated data.

**Table 1.** Data availability by outcome, group, and time intervals.

| Outcome | Serotype (e) | Serostatus (s) | Age (a) | Country (c) | Arm (x) | Months after first dose |
| --- | --- | --- | --- | --- | --- | --- |
| Non-hospitalized case | ✓ | ✓ |  |  | ✓ | [4, 16)<br>[16, 27)<br>[27, 39)<br>[39, 50)<br>[50, 57) |
| Hospitalized case | ✓ | ✓ |  |  | ✓ | [4, 16)<br>[16, 27)<br>[27, 39)<br>[39, 50)<br>[50, 57) |
| Non-hospitalized case |  | ✓ | ✓ |  | ✓ | [4, 16)<br>[16, 27)<br>[27, 39) |
| Hospitalized case |  | ✓ | ✓ |  | ✓ | [4, 16)<br>[16, 27)<br>[27, 39) |
| Non-hospitalized case |  |  |  | ✓ | ✓ | [1, 39) |
| Hospitalized case |  |  |  | ✓ | ✓ | [1, 39) |
| Non-hospitalized and hospitalized cases combined | ✓ |  |  | ✓ | ✓* | [4, 39) |
| Non-hospitalized case | ✓ | ✓ |  | ✓** | ✓ | [4, 57) |
| Hospitalized case | ✓ | ✓ |  | ✓** | ✓ | [4, 57) |

\*Available for placebo arm only
\*\*Available for Sri Lanka only

### Model structure

Since the data were available for multiple time intervals that varied in length, and the size of the population at risk varied over time, we used a survival model for the occurrence of events (i.e., cases and hospitalizations) during the trial. Additionally, the risk of an event in the vaccine arm may have changed over time due to potential waning of the vaccine (16–19,25,28). Using a survival model allowed us to account for these factors. We used this model to estimate the probability of a particular outcome by a given point in time for individuals with a given status, which derives from a cumulative hazard. The cumulative hazard itself is calculated using a hazard rate function, which we describe below.

The core of this approach is the hazard rate function, which we constructed as a function of time and model parameters. Defined as the instantaneous rate at which outcome *o* occurs, where *o* is either a non-hospitalized or hospitalized case, the hazard rate function depends on the force of infection, λ, which is the instantaneous rate at which infection occurs. Relative to the force of infection, the hazard rate function is reduced proportionally by the serotype-specific susceptibility of the individual to infection, π; the probability that an individual becomes infected with serotype *e* before another serotype in country *c*, λ_*c,e*_ / Σ_*e*_ λ_*c,e*_; the serostatus-specific probability of the disease outcome conditional on infection, ρ_*s*_; the country-specific probability that the disease outcome is reported, μ_*c*_; and the outcome-, serostatus-, and serotype-specific relative risk of the outcome due to vaccination, *RR*_*o,s,e*_. We assume that this relative risk takes its most extreme value, ϕ_*o,s,e*_, initially upon vaccination and thereafter wanes at an outcome- and serostatus-specific rate δ_*o,s*_ towards 1. Therefore, the time-varying per-exposure protection is equivalent to 1-*RR*_*o,s,e*_, denoted *PEP*_*o,s,e*_. Consistent with assumptions made in impact projection models for other DENV vaccines, we assume that TAK-003 functions as a “leaky” vaccine, meaning that every individual is partially protected, in contrast to an “all-or-none” vaccine, which completely protects a subset of individuals (34). Further details, including equations, are provided in Supplement 1.1.

The hazard rate function defined above is specified at the most disaggregated level of the data—i.e., data that is available by serotype, baseline serostatus, age, country, and arm. However, as detailed in Table 1, data were not available by all these stratifications simultaneously. In particular, data were aggregated across groups (with respect to serostatus, age, and country). Therefore, to match each piece of data to a corresponding hazard rate function, we derived expected hazard rate functions that took expectations of the disaggregated hazard rates by weighing each of them by the proportional representation of each subgroup within the aggregated group (Supplement 1.2). For example, to obtain a hazard rate function that corresponded to case data broken down by serotype and serostatus, we multiplied the hazard rate *H*_*o_e, g_c,x,s,a*_ by the proportion of participants in each age group, *P*_*a*_, and the proportion of participants in each country, *P*_*c*_, then summed across all age groups and countries such that 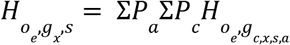.

We used the resulting hazard rate function to calculate the probability of an individual experiencing an outcome within a specified time interval. This event probability is defined as the probability that an individual within a particular group experiences a particular outcome within a particular time interval *i*, provided that they have survived until the previous time interval, *i*-1 (Supplement 1.1). This requires calculating the survival probability, which is a function of the cumulative hazard, or the risk that has been accrued up to time interval *i* (Supplement 1.1). For data that were aggregated by serotype, we calculated the probability of an individual experiencing a non-hospitalized case or hospitalized case by aggregating serotype-specific event probabilities (Supplement 1.2). Similarly, for the situation in which data were not stratified by non-hospitalized and hospitalized cases, but were available by serotype, we aggregated across outcome-specific probabilities (Supplement 1.2).

Once we obtained event probabilities that corresponded to the level of granularity observed for each piece of data, we constructed our likelihood (Supplement 1.2). Each piece of data described in Table 1, for each time interval, was incorporated into this likelihood. For data that were stratified by outcome *o* and serotype *e* within time interval *i*, we assumed the data followed a multinomial distribution characterized by nine possible events, as events can be stratified into four serotypes *e*, two outcome types *o*, and whether an individual survives to the end of time interval *i* (Supplement 1.2). For data that were broken down by outcome *o* but not by serotype *e* within time interval *i*, we assumed the data followed a multinomial distribution with three possible events, as events were broken down into the two outcome types *o* and the event an individual survives to the end of time interval *i* (Supplement 1.2). Finally, we assumed that the number of individuals in each country who were seropositive at baseline followed a binomial distribution (Supplement 1.2).

### Parameter inference

We selected priors for the historical and trial forces of infection such that the mean was approximately equal to the average force of infection estimated by Cattarino et al. (31) for the countries included in the trial. We constructed these priors to have a 95% uncertainty interval of 0.01–0.1, which is wider than was estimated by Cattarino et al. to allow for forces of infection outside the expected range. This was important to account for, since DENV force of infection is known to vary over time (36). For the probabilities of outcomes that depend on serostatus, we selected a distribution such that the mean aligned with estimates of the probability of apparent and hospitalizable disease based on serostatus used by España et al. in impact projection estimates for a previous dengue vaccine (37). As an estimate of variability was not provided, we chose to allow the 95% uncertainty interval to include values above and below the mean by approximately 0.2.

Our priors for per-exposure protection and the duration of protection were purposely constructed to be weakly informative, so that trial data would be the driving force in estimating those quantities. The prior for duration of protection was selected to be a gamma distribution with a mean of 6.7 years and 95% uncertainty interval of 2.2–13.7 years. We selected a prior for per-exposure protection that was centered at zero (i.e., null protection) and had approximately 68% of its weight below zero to account for the potential of the vaccine enhancing the risk of disease, based on a previous dengue vaccine (8). Due to our lack of prior knowledge as to how the probability of reporting a disease outcome might differ by country, we constructed a weakly informative prior for this parameter with a beta prior distribution with mean 0.67 and 95% uncertainty interval of 0.16–0.99.

We used a Hamiltonian Monte Carlo Markov Chain (MCMC) algorithm implemented in RStan (38) in R 4.2.1 (39) to fit the model to trial data and estimate parameter values. We ran four chains with 10,000 iterations and a burn-in period of 2,000 iterations each. We utilized visual inspection of traces and the Gelman-Rubin diagnostic test to assess convergence for all parameters and considered 1.01 to be an acceptable threshold for parameter convergence.

### Model validation

We performed an analysis of our model with simulated data to validate our inference method. To do so, we first sampled 100 sets of parameter values from the priors described above. We used these known parameter values to simulate 100 data sets. We then fitted our model to each of those 100 simulated datasets. We analyzed the frequency of known parameter values being contained within the estimated credible intervals derived from fitting the model, which we refer to as coverage. We also estimate the concordance correlation coefficient for each parameter, which is a quantification of the agreement between known and estimated parameter values (40). Values closer to zero indicate lower agreement, whereas a value of one indicates perfect agreement.

## RESULTS

### Parameter convergence and model fit

All MCMC chains successfully converged, as indicated by Gelman-Rubin diagnostics below 1.01 for all parameters. Additionally, the model generally performed well in fitting to the data, as indicated by the fact that 99% of the data were contained within the respective 95% posterior prediction intervals when aggregated across time intervals (Supplement 2.2). One exception was hospitalization due to DENV-2 in Sri Lanka, which the model underpredicted. In Sri Lanka, rates of hospitalized DENV-2 were unusually high compared to the other serotypes, with this unevenness in hospitalization rates across serotypes not seen in other countries. Therefore, this particular combination of serostatus-specific probability of severe disease and country-specific probability of hospitalization was not identifiable. Additionally, the model occasionally underpredicted or overpredicted case data when disaggregated across time intervals (Supplement 2.4). This was likely due to our assumption that serotype- and country-specific force of infection is constant during the trial. This explanation is supported by the fact that we do not see this same underprediction or overprediction when case data were aggregated across the trial as a whole (Supplement 2.2).

### Model validation

In a validation analysis with simulated data, our model was generally able to recover true values of parameters as indicated by relatively high coverage values across parameters, with a range of 75% to 99% across all parameters (Supplement 2.5). Certain categories of parameters were generally better informed than others. For example, protection against non-hospitalized disease, protection against hospitalized disease, and trial force of infection were better informed compared to historical force of infection and reporting probability. These differences reflect the data that were made available. For example, data on serotype-specific seropositivity by country were not available, so estimates of historical force of infection were largely informed by serotype-specific case data, which also informed estimates of trial force of infection.

Additionally, for some parameters, although coverage estimates were high, the concordance correlation coefficient was relatively low. One such example was protection against both non-hospitalized and hospitalized disease caused by DENV-2 across serostatuses.

However, this is largely because the range from which the true values were sampled was relatively small, as we sampled from posterior distributions from the fitted model. Therefore, numerically small differences between true and estimated values were magnified relative to other parameters that were distributed across a wider range of values.

### Estimates of vaccine parameters

#### Protection against non-hospitalized disease

We estimated that protection against non-hospitalized disease given infection varied considerably by both infecting serotype and baseline serostatus (Fig. 1). Protection upon initial vaccination (captured by parameter ϕ_*d,s,e*_) appeared to be highest against DENV-2, with a median per-exposure protection (PEP) of 99.6% (95% CrI: 96.4, 100.0) among seronegatives and 99.5% (95% CrI: 96.3, 100.0) among seropositives (Fig. 1). In contrast, protection was lower against DENV-3, where it ranged from a median PEP of 26.7% (95% CrI: −8.2, 54.9) among seronegatives to 46.1% (95% CrI: 31.0, 61.1) among seropositives (Fig. 1). PEP was generally higher for seropositive individuals across serotypes – for DENV-1, the probability that protection was higher among seropositive individuals was 99.0%, while this probability was 47.3% for DENV-2, 87.7% for DENV-3, and 93.2% for DENV-4. This is consistent with the published trial results, as efficacy against non-hospitalized disease was higher among seropositive individuals for all serotypes except DENV-2 (25).

**Figure 1.**
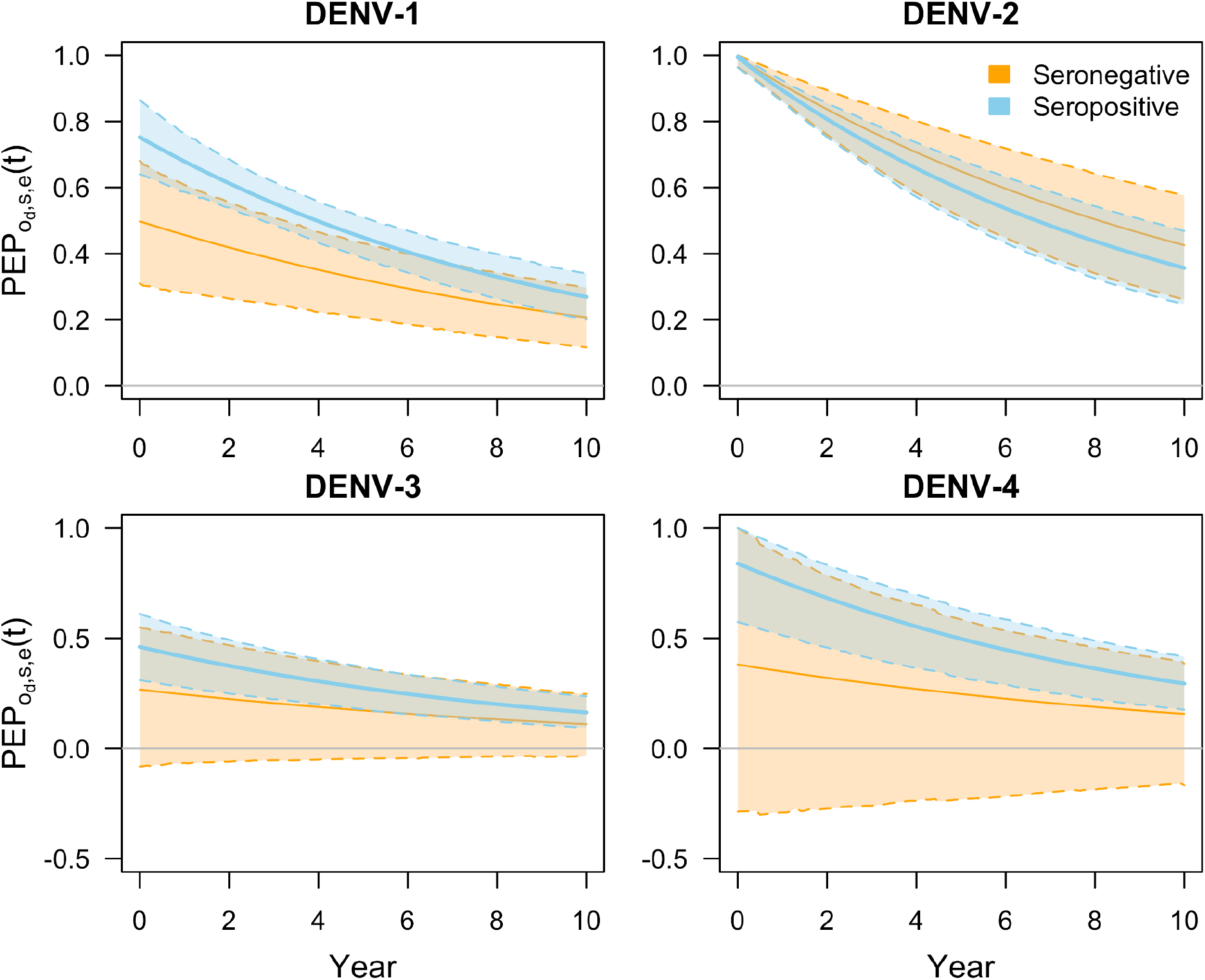
Per-exposure protection against non-hospitalized disease over time, denoted as PEP_o_d,s,e_(t), for each of the four serotypes (panels) by baseline serostatus (colors). The parameters ϕ_o_d,s,e_ occur at time zero, and the waning rate parameter δ_o_d,s,e_ determines how steeply protection changes over time thereafter. Solid lines show median predictions and bands with dashed lines show 95% prediction intervals. A small portion of the low-quantile values of ϕ_o_d,s,e_ are associated with an increase in protection over time, which is a result of the fact that some values for ϕ_o_d,s,e_ were negative and we assume 1-RR_o_d,s,e_(t) approaches 0 as time passes.

We also observed the impact of defining a separate duration of protection for seronegative and seropositive individuals. We estimated a slightly shorter duration of protection for seropositive individuals (9.8 years [95% CrI: 7.3, 13.8]) compared to seronegative individuals (11.8 years [95% CrI: 7.8, 19.6]), with a 74.2% probability that duration of protection is shorter for seropositive individuals. This effect is clear for an example such as protection against DENV-2 (Fig. 1), because initial protection across serostatuses is similar, with an initial protection of 99.6% (95% CrI: 96.4, 100.0) for seropositive individuals and 99.5% (95% CrI: 96.3, 100.0) for seronegative individuals. After ten years, per-exposure protection decreased to 35.7% (95% CrI: 24.6, 46.9) for seropositive individuals, but to 42.6% (95% CrI: 26.1, 57.6) for seronegative individuals. We also note that it appears as though the credible interval for per-exposure protection narrows for protection against DENV-3 and DENV-4. However, this is because posterior estimates that were negative at the time of vaccination increase towards zero while positive estimates decrease towards zero over time, as a result of our choice of mathematical function for how protection changes over time. This affects only a small portion of our posterior estimates.

#### Protection against hospitalized disease

Our estimates of initial per-exposure protection conferred by the vaccine against hospitalized disease (captured by parameter ϕ_*h,s,e*_) followed similar patterns as our estimates of protection against non-hospitalized disease. Namely, we observed differences in protection by serotype and serostatus (Fig. 2). Similarly, protection against hospitalized disease due to DENV-2 was the highest of the four serotypes, particularly among seropositives (98.8% [95% CrI: 90.1, 100.0]). On the other hand, protection against hospitalized disease resulting from DENV-3 ranged from −72.7% (95% CrI: −149.4, −4.0) among seronegatives to 52.3% (95% CrI: 10.9, 88.4) among seropositives. PEP against hospitalized disease was higher among seropositive individuals infected with DENV-2 and DENV-3, with probabilities of 57.6% and 99.9% respectively, but was higher among seronegative individuals infected with DENV-1 and DENV-4, with probabilities of 96.8% and 51.3% respectively. These trends are consistent with previously published efficacy estimates for DENV-1 and DENV-3 (25). For DENV-2, previously published estimates indicate higher protection among seronegatives, though this is based on a one-sided confidence interval due to zero cases in the vaccine arm (25). For DENV-4, efficacy could not be calculated due to insufficient case counts (25).

**Figure 2.**
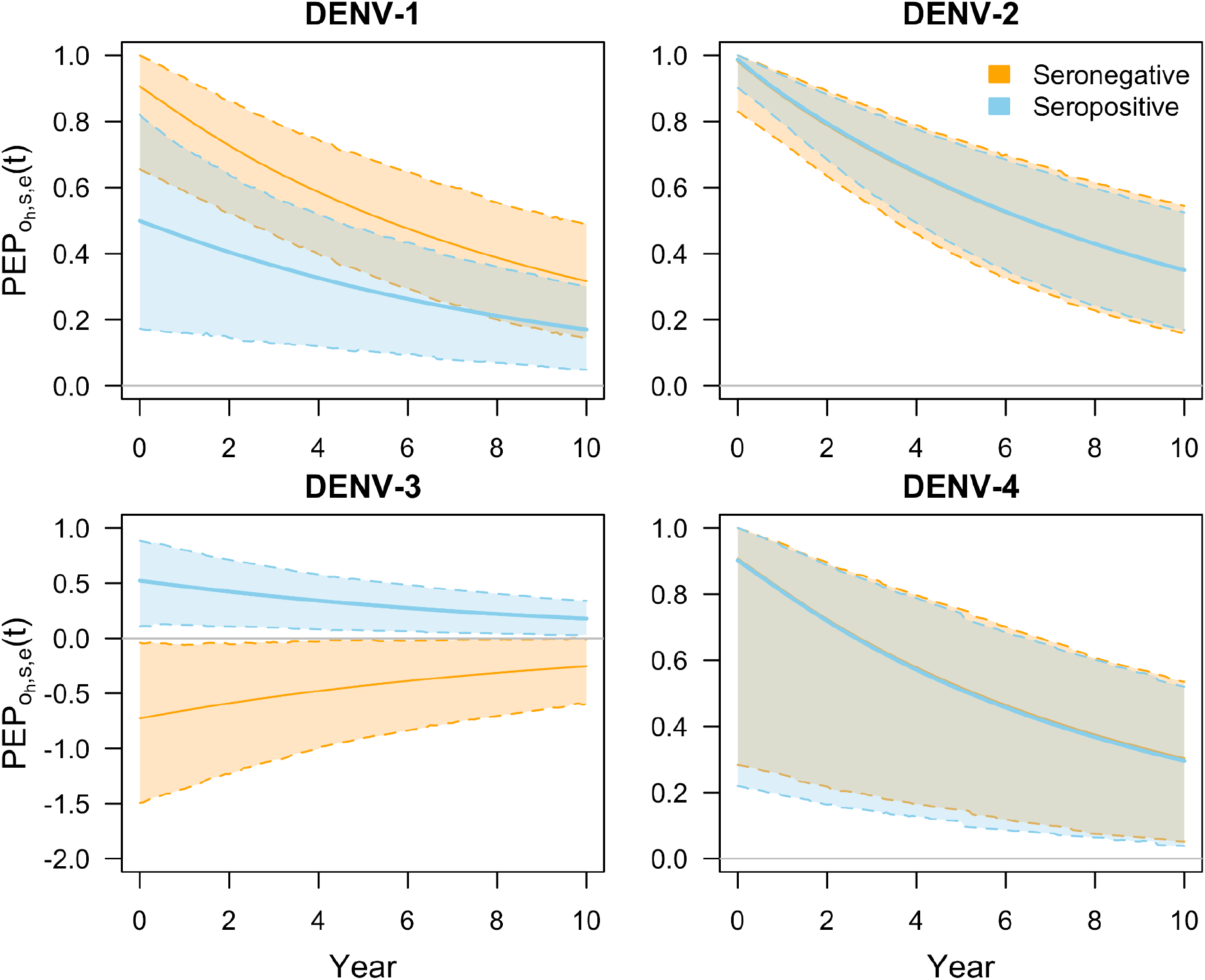
Per-exposure protection against hospitalized disease over time, PEP_o_h,s,e_(t), for each of the four serotypes (panels) by baseline serostatus (colors). The parameters ϕ_o_h,s,e_ occur at time zero, and the waning rate parameter δ_o_h,s,e_ determines how steeply protection changes over time thereafter. Solid lines show median predictions and bands with dashed lines show 95% prediction intervals. For seronegatives infected with DENV-3, values of ϕ_o_h,s,e_ are associated with an increase in protection over time, which is a result of the fact that the values for ϕ_o_h,s,e_ were negative and we assume 1-RR_o_h,s,e_(t) approaches 0 as time passes.

Our estimates of per-exposure protection against hospitalized disease after a ten-year period reflect the fact that we estimated a very similar duration of protection for seronegative (9.93, 95% CrI: [6.0, 19.1]) and seropositive (9.78, 95% CrI: [6.0, 17.3]) individuals (Fig. 2). For example, for protection against DENV-2, initial protection for seronegatives was 98.0% (95% CrI: 83.0, 100.0) and waned to 34.9% (95% CrI: 15.8, 54.4). Similarly, initial protection for seropositives was 98.8% (95% CrI: 90.1, 100.0) and waned to 35.0% (95% CrI: 16.9, 52.5). Additionally, protection against DENV-3 appeared to increase over time for baseline seronegative individuals, from −72.7% (95% CrI: −149.4, −4.0) to −25.4% (95% CrI: −59.9, −1.1), due to our assumption that relative risk approaches one in the long term regardless of its initial value.

#### Estimating vaccine efficacy assuming independent outcome probabilities

To illustrate why our estimates of protection for seronegatives infected with DENV-3 and DENV-4 were less uncertain than those reported by Takeda, we constructed a simplified version of our model that follows the assumption that the baseline probabilities for each outcome are independent by serotype, serostatus, and outcome type (hereafter referred to as the “independent-parameter model”). This independent-parameter model reflects the assumptions made by Takeda in their calculations of vaccine efficacy (16–19,25). In comparison, our full model accounts for interdependencies by sharing parameters across baseline probabilities. We fitted the independent-parameter model to cumulative trial data on non-hospitalized and hospitalized cases by infecting serotype and baseline serostatus, which allowed us to obtain serotype- and serostatus-specific estimates of efficacy for comparison against our full model’s estimates and those made by Takeda.

Estimates of vaccine efficacy from the independent-parameter model very closely matched vaccine efficacy estimates by Takeda, for both non-hospitalized (Fig. 3, top) and hospitalized disease (Fig. 3, bottom). In particular, the estimates from this model match the relatively wide uncertainty reported previously for the serotype and serostatus combinations of interest, unlike the per-exposure protection estimates from the full model, which are also shown.

**Figure 3.**
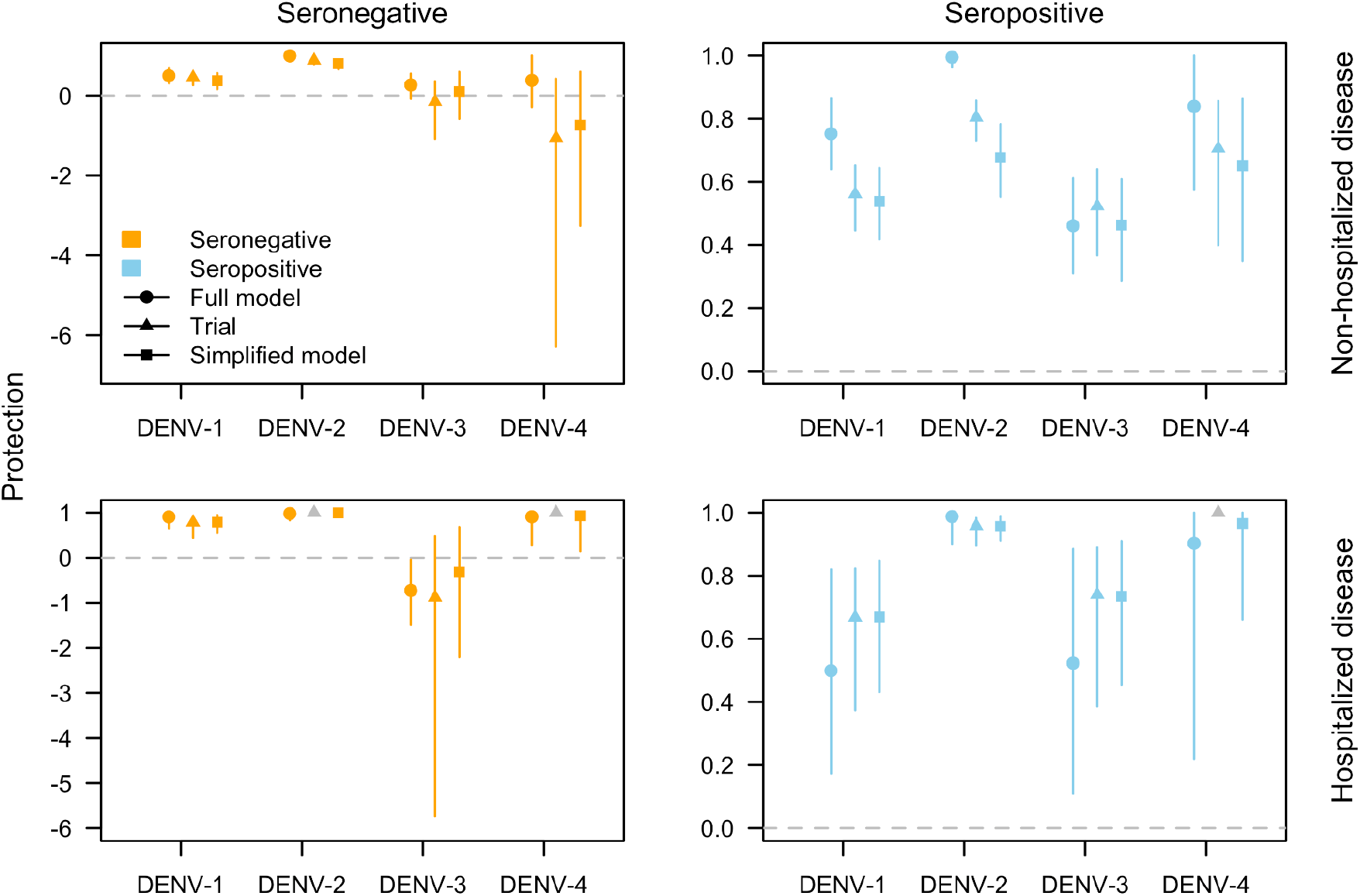
Estimates of protection conferred by the vaccine as per-exposure protection estimated by the full model (circle), efficacy reported by the trial (triangle), and efficacy estimated with the simplified independent-parameter model (square). Lines indicate 95% uncertainty intervals. Grey shapes indicate an estimate could not be reported. Protection is broken down by infecting serotype, non-hospitalized (top row) or hospitalized disease (bottom row), and baseline serostatus (left column for seronegative, right column for seropositive).

For example, the independent-parameter model estimated a vaccine efficacy for non-hospitalized disease among seronegatives infected with DENV-4 of −74.2% (95% CrI: −326.2, 59.5) (cf. −105.6%; 95% CI: −628.7, 42.0 from (41)). For hospitalized disease among seronegatives infected with DENV-3, the independent-parameter model estimated a vaccine efficacy of −32.0% (95% CrI: −220.8, 68.0) (cf. −87.9%; 95% CrI: −573.4, 47.6 from (25)).

The results from the independent-parameter model show the impact of not accounting for interdependent epidemiological processes when estimating stratum-specific vaccine parameters. For example, in our full model, we account for the fact that individuals of the same serostatus share both a probability of disease given infection and a probability of hospitalization given disease. Our full model also explicitly accounts for the fact that hospitalization is conditional on developing symptomatic disease, allowing data from non-hospitalized and hospitalized cases to jointly inform these parameters. In addition, by including the probability that an individual encounters a particular serotype before the other three in our hazard, we allow data for all four serotypes within the same country to inform estimates of each serotype-specific force of infection, even when fitting to data pertaining to a single serotype. All these assumptions reflect epidemiological realities more closely than the relatively strong assumptions made in the original analysis of these data. By accounting for these interdependencies, we can leverage data across multiple strata to jointly inform model parameters, which helps reduce parameter uncertainty. This effect is especially useful when observed endpoint events are low, such as for DENV-4 in the TAK-003 efficacy trial. Additionally, it is important to note that our full model is estimating per-exposure protection, rather than efficacy, which differs somewhat in its interpretation. This may also explain some of the observed discrepancy between our model’s results and those reported by the trial and estimated with our independent-parameter model, both of which are estimating efficacy.

## DISCUSSION

We fitted a multi-level model to publicly available trial data to estimate individual-level protection parameters for the TAK-003 dengue vaccine. Our model structure combined hazard rates across multiple aggregation levels, allowing us to leverage and pool disparate data to inform parameters. In doing so, we used all published epidemiological data, including country-level data, which has not yet been done in previous analyses (16–19,25,28). By using this approach, we have enhanced understanding of the characteristics of TAK-003, in particular by reducing uncertainty around estimates of serotype-specific protection. In addition, unlike reported efficacy estimates, we quantified per-exposure protection, which is more appropriate for use in modeling projections of public health impact and cost-effectiveness.

Using this approach, we reduced uncertainty around estimates of individual-level protection against hospitalized disease due to DENV-3 among seronegative individuals compared to reported efficacy estimates, and estimated protection against non-hospitalized disease among seronegatives due to both DENV-3 and DENV-4 to be more tightly distributed near the null. For example, while the median of our posterior estimate of protection against hospitalized disease among seronegatives due to DENV-3 was quite similar to the mean estimate of reported vaccine efficacy (−72.7% and −87.9%, respectively), the range of our 95% credible interval was 149.4 while the range of the 95% confidence interval reported in the trial was 621. We can attribute this reduced uncertainty to the fact that we pooled data across serotypes, serostatus, and outcomes to jointly inform parameter estimates. To confirm this hypothesis, we developed a simplified model in which we estimated the probability of an individual experiencing an outcome independently by serotype, serostatus, and outcome type. Protection estimated by this model matched the relatively wide uncertainty observed in the trial estimates. Jointly leveraging the data across stratifications—such as serotype, serostatus, country, and outcome type—reflects our assumption that aspects of the disease process—such as baseline probabilities of disease and reporting probabilities—are shared across some of these stratifications. This assumption is more epidemiologically sound than assuming that the probabilities of each outcome are independent. Our results indicate that accounting for these dependencies is particularly useful for obtaining more precise estimates of protection for endpoints with low case counts, such as DENV-4 in this trial.

Other advantages of our modeling approach include the fact that we built in the ability to combine hazards across levels of stratification, which we used to estimate cumulative hazards for each time interval for which data were available. For each time interval, trial data were available at different levels of aggregation, so combining hazards to match the aggregation of the data enabled us to integrate all publicly available data into our model. For example, at the time interval of 4-16 months, data were available on a serostatus- and serotype-specific level and separately on age- and serostatus-specific level. Additionally, using a survival model allowed us to leverage all time intervals for which data were available. This was particularly important because some data types were only available at certain time intervals. Leveraging all the data across varying stratifications and temporal availability gave us the ability to estimate parameters at finer aggregation levels than were present in much of the data. For example, the serotype- and country-specific case data were only available for a single time interval and a single arm. Because we were able to both aggregate our hazard rates and calculate cumulative hazards for each time interval, we were able to incorporate all data in our model to inform our estimates of country- and serotype-specific forces of infection during the trial.

There are also some notable limitations of our approach. One is that our study does not provide mechanistic insight as to why we see differential protection by serotype, serostatus, and outcome. For example, we did not explicitly rule out the possibility that TAK-003 acts as a silent infection, like Dengvaxia, in which vaccinated seronegative individuals are at higher risk of severe disease upon breakthrough infection (8). However, this does not appear likely given that our results indicate that any possible enhanced risk of severe disease is serotype-specific, namely to individuals infected with DENV-3 or DENV-4, whereas the enhanced risk of severe disease for a natural secondary infection is not typically thought to be specific to the order of infecting serotypes. Another possibility is that TAK-003 modulates the risk of disease by inducing differential levels of serotype-specific antibody titers that decay over time (42–44).

Other recent work has explored this hypothesis with an alternative model of TAK-003 protection that includes latent variables for antibody titer dynamics (28). Unlike this study, however, we estimated country-specific forces of infection averaged over the entire trial, rather than time-varying forces of infection. We felt that allowing for country-specific forces of infection was important for our analysis given its emphasis on the interdependencies across different publicly available aggregations of data. Another limitation of our approach is that we relied on the trial’s definition of hospitalized disease as a proxy for severe disease, which can be problematic due to variability in clinical practices and definitions across geographical settings (12,32). This is a limitation for all analyses of these data, however. We at least sought to address this problem by including country-specific reporting probabilities by outcome in our model. Relatedly, we did not estimate protection against infection due to the fact that data on subclinical infections were not collected as part of the trial (45).

In summary, we developed an approach to leverage clinical trial data over a range of levels of aggregation to estimate direct protection offered by a novel dengue vaccine. We found that the vaccine induces differential protection by infecting serotype, baseline serostatus, and outcome, consistent with previous work (16–19,25,28). Although our estimates of per-exposure protection are more precise than previously reported estimates of efficacy, we still cannot rule out the possibility of enhanced risk of disease upon vaccination. This uncertainty can be resolved with additional endpoint events caused by DENV-3 and DENV-4, which are being pursued through ongoing trials being conducted by Takeda (WHO) (46). Our approach will remain useful in enhancing the interpretation of those trials given its ability to account for innate interdependencies among population strata, which, as we have demonstrated, have major implications for quantifying uncertainty about vaccine protection.

## Data Availability

All data used are published as cited in the manuscript.

## Acknowledgements

This work was supported by the NIH National Institute of General Medical Sciences R35 MIRA program to T.A.P. (grant no. R35GM143029) and by a contract from the World Health Organization to the University of Notre Dame.

