## Supplement for "Refining uncertainty about the TAK-003 dengue vaccine with a multi-level model of clinical efficacy trial data"

Manar Alkuzweny, Guido España, T. Alex Perkins
Department of Biological Sciences and Eck Institute of Global Health, University of Notre Dame

### Table of contents

**1. Supplemental Methods** **2**
- 1.1 General analytical framework 2
- 1.2 Adaptation of the general analytical framework to available data 6
- 1.3 Derived parameters 11
- 1.4 Independent-parameter model 12

**2. Supplemental Results** **14**
- 2.1 Estimates of per-exposure protection 14
- 2.2 Model predictions compared to trial data by data type 15
- 2.3 Epidemiological parameter estimates 22
- 2.4 Model predictions broken down by time intervals 28
- 2.5 Model validation results 38

### 1. Supplemental Methods

1.1 General analytical framework

The data from the trial consist of disease outcomes, *o*, reported within time intervals $[t_{i-1},t_i)$. There are eight disease outcomes, $o_{d,e}$ and $o_{h,e}$ for each of $e\in\{1,2,3,4\}$, where *d* refers to non-hospitalized disease, *h* refers to hospitalized disease, and *e* refers to serotype. When an outcome occurs during this interval, we can say that its timing, $t_o$, satisfies $t_{i-1}\le t_o<t_i$. These outcomes are reported separately for different groups, *g*, within the study population. We assume that these outcomes occur according to a hazard rate function, $H_{o,g}(t,\boldsymbol{\theta})$, which is a function of time and a vector of model parameters, $\boldsymbol{\theta}$. At time $t_{i-1}$, $N_{g,i}$ individuals in group *g* have the potential to experience the outcome. By time $t_i$, $O_{o,g,i}$ of those $N_{g,i}$ individuals experience the outcome. Over time, the number who were potentially at risk at the start of the trial, $N_{g,0}$, is decremented by each $O_{o,g,j}$ for all $j<i$ and for all $o\in\{o\}$ to obtain

$$N_{g,i} = N_{g,0} - \sum_{o}\sum_{j<i} O_{o,g,j}. \quad (1)$$

The eight possible disease outcomes are mutually exclusive. As such, the contribution to the likelihood from individuals in group *g* during interval *i* is equal to the probability that the $N_{g,i}$ individuals who were potentially at risk at time $t_{i-1}$ experience outcomes by time $t_i$ that are distributed as

$$\left\{O_{o_{d,1},g,i}, O_{o_{d,2},g,i}, O_{o_{d,3},g,i}, O_{o_{d,4},g,i}, O_{o_{h,1},g,i}, O_{o_{h,2},g,i}, O_{o_{h,3},g,i}, O_{o_{h,4},g,i}, O_{o_{\emptyset},g,i}\right\}, \quad (2)$$

where

$$O_{o_{\emptyset},g,i} = N_{g,i} - \sum_{e}\left(O_{o_{d,e},g,i} + O_{o_{h,e},g,i}\right) \quad (3)$$

is the number of individuals who experience the null outcome, $o_{\emptyset}$, in which no disease outcome is observed. The probability of the set of outcomes in eqn. (2) is described by a multinomial distribution with probabilities for each individual outcome *o* of

$$\Pr(o|g,i) = \frac{S\left(t_{i-1}|o,g,\boldsymbol{\theta}\right)-S\left(t_i|o,g,\boldsymbol{\theta}\right)}{S\left(t_{i-1}|\{o\},g,\boldsymbol{\theta}\right)} \quad (4)$$

for the eight disease outcomes and

$$\Pr\left(o_{\emptyset}|g,i\right) = 1 - \sum_{e}\left(\Pr\left(o_{d,e}|g,i\right) + \Pr\left(o_{h,e}|g,i\right)\right) \quad (5)$$

for the null outcome. The survival probability is defined as

$$S(t|o,g,\boldsymbol{\theta}) = \exp\left(-\int_0^t H_{o,g}(t,\boldsymbol{\theta})dt\right), \quad (6)$$

which yields the probability that outcome *o* has not occurred by time *t*. Relatedly,

$$S(t|\{o\},g,\boldsymbol{\theta}) = 1 - \sum_{o}(1 - S(t|o,g,\boldsymbol{\theta})) \quad (7)$$

is the probability that an individual has not experienced any disease outcome by time *t*.

Defined as the instantaneous rate at which outcome *o* occurs, the core of the hazard rate function is the force of infection, λ, which is the instantaneous rate at which infection occurs. Relative to the force of infection, the hazard rate function is reduced proportionally by the susceptibility of the individual to infection, π; the probability that an individual becomes infected with serotype *e* before another serotype, $\lambda_{c,e} / \Sigma_e \lambda_{c,e}$; the serostatus-specific probability of the disease outcome conditional on infection, $\rho_s$; the country-specific probability that the disease outcome is reported, $\mu_c$; and the outcome-, serostatus- and serotype-specific relative risk of the outcome due to vaccination, $RR_{o,s,e}$. We assume that this relative risk takes its most extreme value, $\phi_{o,s,e}$, initially upon vaccination and thereafter wanes at an outcome- and serostatus-specific rate $\delta_{o,s}$ towards 1. Consistent with assumptions made in impact projection models for other DENV vaccines, we assume that TAK-003 functions as a "leaky" vaccine, meaning that every individual is partially protected, in contrast to an "all-or-none" vaccine, which completely protects a subset of individuals (34-35,37). See Tables S1 and S2 for full definitions of mathematical symbols used.

In an idealized case in which data on groups were available in the most disaggregated way possible—i.e., simultaneously by age group, arm, country, serostatus, and serotype—the hazard rate function for non-hospitalized disease with serotype *e*, $o_{d,e}$, would be

$$H_{o_{d,e}, g_{c,x,s,a}}(t, \boldsymbol{\theta}) = \pi_{c,e,s,a} \lambda_{c,e} \frac{\lambda_{c,e}}{\sum_e \lambda_{c,e}} \rho_{o_d,s} RR_{o_d,x}(t, \phi_{o_d,s,e}, \delta_{o_d,s})(1 - \rho_{o_h,s} RR_{o_h,x}(t, \phi_{o_h,s,e}, \delta_{o_h,s}))\mu_{o_d,c} \quad (8)$$

for individuals in group $g_{c,x,s,a}$. Similarly, the hazard rate function for hospitalized disease with serotype *e*, $o_{h,e}$, would be

$$H_{o_{h,e}, g_{c,x,s,a}}(t, \boldsymbol{\theta}) = \pi_{c,e,s,a} \lambda_{c,e} \frac{\lambda_{c,e}}{\sum_e \lambda_{c,e}} \rho_{o_d,s} RR_{o_d,x}(t, \phi_{o_d,s,e}, \delta_{o_d,s}) \rho_{o_h,s} RR_{o_h,x}(t, \phi_{o_h,s,e}, \delta_{o_h,s}) \mu_{o_h,c} \quad . \quad (9)$$

In both eqns. (8) and (9), $RR_{o,x}$=1 for unvaccinated individuals, denoted $x$=$x_u$.

***Table S1.*** *Mathematical symbols other than model parameters, which are defined in Table S2.*

| Symbol | Definition |
|---|---|
| $N$ | Number of trial participants potentially at risk of an outcome |
| $O$ | Number of trial participants experiencing an outcome |
| $t$ | Time |
| $i$ | Time interval spanning $[t_{i-1},t_i)$ |
| $o$ | Index for a type of outcome, the collection of which is denoted $\{o\}$ and includes all $o_{d,e}$ and $o_{h,e}$ |
| $d$ | Denotes an outcome of non-hospitalized disease |
| $h$ | Denotes an outcome of hospitalized disease |
| $e$ | Denotes a dengue virus serotype, which span {1,2,3,4} |
| $g$ | Index for a group of individuals, the collection of which is denoted $\{g\}$ and includes all $g_{c,x,s,k,a}$ |
| $c$ | Denotes a country in which trial participants reside |
| $x$ | Denotes the vaccination status of trial participants, with $x_v$ denoting vaccinated individuals and $x_u$ denoting unvaccinated individuals |
| $s$ | Serostatus, with $s_p$ denoting seropositive individuals and $s_n$ denoting seronegative individuals |
| $a$ | Denotes an age group defined by minimum age $a_1$ and maximum age $a_2$ |
| $p$ | Proportion of trial participants in a given group $g$ |
| $H$ | Hazard rate function |
| $S$ | Survival probability |
| $RR$ | Relative risk of a disease outcome, which depends on the type of outcome $o$ and vaccination status $x$ |

***Table S2.*** *Mathematical symbols for model parameters, with other symbols defined in Table S1.*

| Symbol | Definition | Parameter type | Prior |
|---|---|---|---|
| $\bar{\lambda}$ | Force of infection prior to the trial, which depends on country *c* and serotype *e* | Estimated | $log(\bar{\lambda})$ ~ Normal(-3.4, 0.6) |
| π | Probability that a trial participant is susceptible at baseline, which depends on country *c*, serotype *e*, serostatus *s*, and age group *a* | Derived | N/A |
| λ | Force of infection during the trial, which depends on country *c* and serotype *e* | Estimated | $log(\lambda)$ ~ Normal(-3.4, 0.6) |
| ρ | Probability of an outcome for an unvaccinated person, which depends on the type of outcome *o* and serostatus *s* | Estimated | $\rho_{d,s\_n}$ ~ Beta(6, 14)<br>$\rho_{d,s\_p}$ ~ Beta(12, 8)<br>$\rho_{h,s\_n}$ ~ Beta(2.2, 17.8)<br>$\rho_{h,s\_p}$ ~ Beta(4, 16) |
| μ | Probability that a disease outcome is reported, which depends on the type of outcome *o* and the country *c* | Estimated | μ ~ Beta(2, 1) |
| ϕ | Initial level of protection against a disease outcome upon vaccination, which represents the most extreme modification of risk due to vaccination and depends on the type of outcome *o*, serostatus *s*, and serotype *e* | Estimated | 1-ϕ ~ Gamma(0.5, 0.5) |
| δ | Rate of waning of the protection against a disease outcome some time after vaccination, which depends on the type of outcome *o* and serostatus *s* | Estimated | δ ~ 1/Gamma(5, 0.75) |

#### 1.2 Adaptation of the general analytical framework to available data

Fully disaggregated data for all possible groups $g_{c,x,s,a}$ were not available to us. Therefore, we derived separate contributions to the likelihood appropriate to all groups for which data were available (Table S3). In the case where outcomes $o$ were reported in an aggregated fashion, we collapsed the set of outcomes in eqn. (2) down to those that were reported (i.e., $\{o_d, o_h, N_{g,i} - o_d - o_h\}$) and the outcome-specific probabilities in eqn. (4) were summed accordingly. In the case where data for groups $g$ were reported in an aggregated fashion, we derived expected hazard rate functions that took expectations of the more disaggregated hazards in eqns. (8) and (9) weighted by the proportional representation of each subgroup within the aggregated group. Below, we explicate the contributions to the likelihood from each of the six forms of aggregated data listed in Table S3. We also provide a schematic of how parameters relate to the available data with Figure S1.

***Table S3.*** *Data aggregations that are publicly available and used in this analysis.*

| **Outcomes** | **Group** | **Intervals** | **Description** |
|---|---|---|---|
| $o_{d,e}$, $o_{h,e}$ | $g_{x,s}$ | [4, 16)<br>[16, 27)<br>[27, 39)<br>[39, 50)<br>[50, 57) | Non-hospitalized and hospitalized disease by serotype ($o_{d,e}$, $o_{h,e}$) stratified by vaccination status and serostatus ($g_{x,s}$) |
| $o_d$, $o_h$ | $g_{x,s,a}$ | [4, 16)<br>[16, 27)<br>[27, 39) | Non-hospitalized and hospitalized disease ($o_d$, $o_h$) stratified by vaccination status, serostatus, and age ($g_{x,s,a}$) |
| $o_d$, $o_h$ | $g_{c,x}$ | [1, 39) | Non-hospitalized and hospitalized disease ($o_d$, $o_h$) stratified by country and vaccination status ($g_{c,x}$) |
| $o_e$ | $g_{c,x\_u}$ | [4, 39) | Disease by serotype ($o_e$) stratified by country for unvaccinated individuals ($g_{c,x\_u}$) |
| $o_{d,e}$, $o_{h,e}$ | $g_{c\_LKA,x,s}$ | [4, 57) | Non-hospitalized and hospitalized disease by serotype ($o_{d,e}$, $o_{h,e}$) for Sri Lanka stratified by serostatus and vaccination status ($g_{c\_LKA,x,s}$) |
| $s$ | $g_c$ | $(-\infty,0)$ | Serostatus at baseline ($s$) stratified by country ($g_c$) |

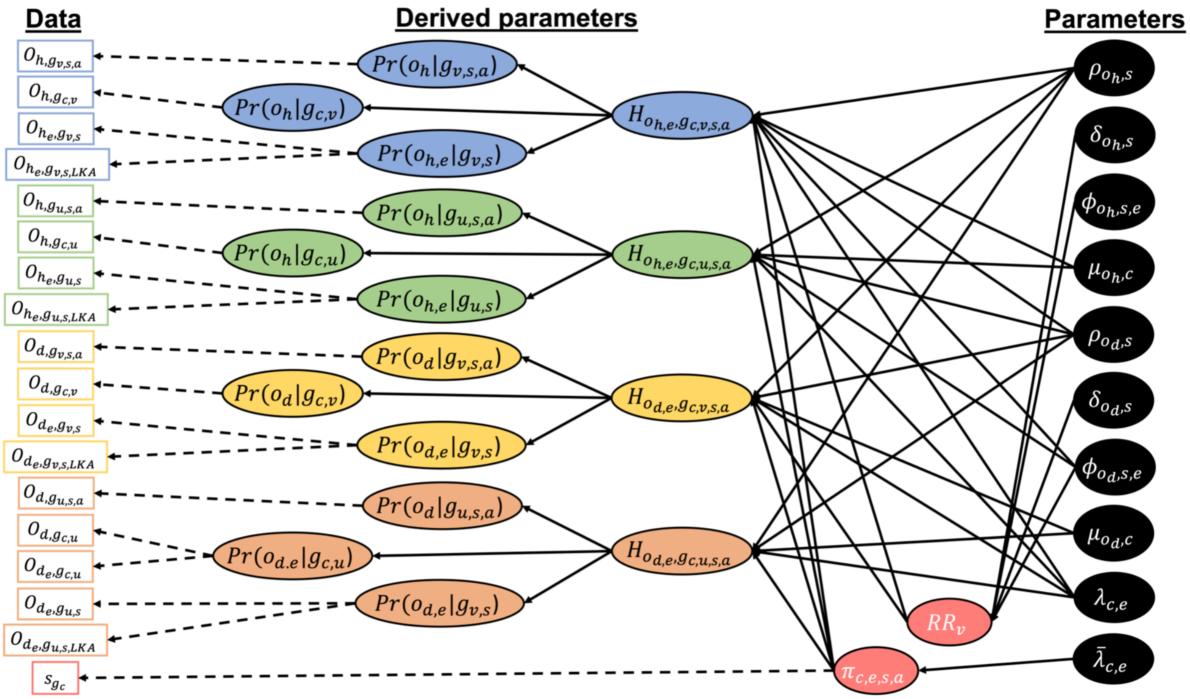


***Figure S1. Relationships among parameters and data in our model.*** *Estimated parameters (black circles) are used to calculate derived parameters (colored circles), which represent the probability of an outcome (o) occurring (d: non-hospitalized disease; h: hospitalized disease; e: serotype) given information on which group (g) a participant belongs to (a: age; x: arm, u for control and v for vaccine; c: country; s: serostatus). Derived parameters are referenced in the likelihood. Solid arrows represent deterministic relationships and dashed arrows represent stochastic relationships. See Table S1 for complete definitions of mathematical symbols.*

*Likelihood for outcomes $o_{d,e}$, $o_{h,e}$ from group $g_{x,s}$*
These data aggregate over country *c* and age *a*. The expected hazard rate functions for this group are then

$$H_{o_{d,e},g_{x,s}}(t,\boldsymbol{\theta}) = \sum_c p_c \sum_a p_a H_{o_{d,e},g_{c,x,s,a}}(t,\boldsymbol{\theta}) \quad (10)$$

and

$$H_{o_{h,e},g_{x,s}}(t,\boldsymbol{\theta}) = \sum_c p_c \sum_a p_a H_{o_{h,e},g_{c,x,s,a}}(t,\boldsymbol{\theta}), \quad (11)$$

where $p_c$ and $p_a$ are the proportions of trial participants in country *c* and age group *a*, respectively. Hence, $\Sigma_c\, p_c = 1$ and $\Sigma_a\, p_a = 1$. Due to a lack of information about how the age distribution of trial participants might have varied across countries, we assumed that the overall age distribution reported for the trial as a whole applied to all countries.

The contribution to the likelihood from this portion of the data is then

$$L_{\{o_{d,e},o_{h,e}\},g_{x,s}}(\boldsymbol{\theta}) = \prod_i \prod_x \prod_s Multinomial\left(\{O_{o,g_{x,s},i}\} \mid \{\Pr(o|g_{x,s},i)\}\right), \quad (12)$$

where $\{O_{o,g_{x,s},i}\}$ includes all nine outcomes as in eqn. (2) for group $g_{x,s}$ and $\Pr(o|g_{x,s},i)$ follows eqns. (4) and (5). The only difference is that the hazard rate functions from eqns. (10) and (11) are used in the calculation of the survival probabilities in eqns. (6) and (7).

*Likelihood for outcomes $o_d$, $o_h$ from group $g_{x,s,a}$*

These data aggregate over serotypes *e* and countries *c*. The expected hazard rate functions for this group are then

$$H_{o_{d,e},g_{x,s,a}}(t,\boldsymbol{\theta}) = \sum_c p_c H_{o_{d,e},g_{c,x,s,a}}(t,\boldsymbol{\theta}) \quad (13)$$

and

$$H_{o_{h,e},g_{x,s,a}}(t,\boldsymbol{\theta}) = \sum_c p_c H_{o_{h,e},g_{c,x,s,a}}(t,\boldsymbol{\theta}) \quad (14)$$

where $p_c$ is the proportion of trial participants in country *c*. Hence, $\Sigma_c\, p_c = 1$.

Because the outcomes reported are aggregated across serotypes, the observed outcomes are $o_d = o_{d,1} \cup o_{d,2} \cup o_{d,3} \cup o_{d,4}$, $o_h = o_{h,1} \cup o_{h,2} \cup o_{h,3} \cup o_{h,4}$, and $o_\varnothing = N_{g,i} - \{o_d \cup o_h\}$. The probabilities corresponding to each of these outcomes are

$$\Pr\left(o_d|g_{x,s,a}, i\right) = \sum_e \Pr\left(o_{d,e}|g_{x,s,a}, i\right), \quad (15)$$

$$\Pr\left(o_h|g_{x,s,a}, i\right) = \sum_e \Pr\left(o_{h,e}|g_{x,s,a}, i\right), \quad (16)$$

and

$$\Pr\left(o_\emptyset|g_{x,s,a}, i\right) = 1 - \Pr\left(o_d|g_{x,s,a}, i\right) - \Pr\left(o_h|g_{x,s,a}, i\right). \quad (17)$$

The probabilities on the right-hand sides of eqns. (15) and (16) follow eqn. (4). The only difference is that the hazard rate functions from eqns. (13) and (14) are used in the calculation of the survival probabilities in eqns. (6) and (7).

The contribution to the likelihood from this portion of the data is then

$$L_{\{o_d,o_h\},g_{x,s,a}}(\boldsymbol{\theta}) = \prod_i \prod_x \prod_s \prod_a Multinomial\left(\{O_{o,g_{x,s,a},i}\} \mid \{\Pr(o|g_{x,s,a}, i)\}\right), \quad (18)$$

where $\{O_{o,g_{x,s,a},i}\} = \{O_{o_d,g_{x,s,a},i}, O_{o_h,g_{x,s,a},i}, O_{o_\emptyset,g_{x,s,a},i}\}$ and $\{\Pr(o|g_{x,s,a}, i)\}$ is a vector of probabilities defined in eqns. (15)-(17).

*Likelihood for outcomes $o_d$, $o_h$ from group $g_{c,x}$*

These data aggregate over serotypes *e*, serostatus *s*, and age group *a.* The expected hazard rate functions for this group are then

$$H_{o_{d,e},g_{c,x}}(t,\boldsymbol{\theta}) = \sum_a p_a \sum_s p_{s,a} H_{o_{d,e},g_{c,x,s,a}}(t,\boldsymbol{\theta}) \quad (19)$$

and

$$H_{o_{h,e},g_{c,x}}(t,\boldsymbol{\theta}) = \sum_a p_a \sum_s p_{s,a} H_{o_{h,e},g_{c,x,s,a}}(t,\boldsymbol{\theta}), \quad (20)$$

where $p_a$ is the proportion of all trial participants in age group $a$ and $p_{s,a}$ is the proportion of trial participants within age group $a$ that have serostatus $s$. Hence, $\Sigma_a p_a = 1$ and $\Sigma_s p_{s,a} = 1$. Due to a lack of information about how the age distribution of trial participants might have varied across countries, we assumed that the overall age distribution reported for the trial as a whole applied to all countries.

Because the outcomes reported are aggregated across serotypes, the observed outcomes are $o_d = o_{d,1} \cup o_{d,2} \cup o_{d,3} \cup o_{d,4}$, $o_h = o_{h,1} \cup o_{h,2} \cup o_{h,3} \cup o_{h,4}$, and $o_\varnothing = N_{g,i} - \{o_d \cup o_h\}$. The probabilities corresponding to each of these outcomes are

$$\Pr\left(o_d | g_{c,x}, i\right) = \sum_e \Pr\left(o_{d,e} | g_{c,x}, i\right), \quad (21)$$

$$\Pr\left(o_h | g_{c,x}, i\right) = \sum_e \Pr\left(o_{h,e} | g_{c,x}, i\right), \quad (22)$$

and

$$\Pr\left(o_\emptyset | g_{c,x}, i\right) = 1 - \Pr\left(o_d | g_{c,x}, i\right) - \Pr\left(o_h | g_{c,x}, i\right). \quad (23)$$

The probabilities on the right-hand sides of eqns. (21) and (22) follow eqn. (4). The only difference is that the hazard rate functions from eqns. (19) and (20) are used in the calculation of the survival probabilities in eqns. (6) and (7).

The contribution to the likelihood from this portion of the data is then

$$L_{\{o_d, o_h\}, g_{c,x}}(\boldsymbol{\theta}) = \prod_i \prod_c \prod_x Multinomial\left(\{O_{o, g_{c,x}, i}\} \mid \{\Pr(o | g_{c,x}, i)\}\right), \quad (24)$$

where $\{O_{o,g_{c,x},i}\} = \{O_{o_d,g_{c,x},i}, O_{o_h,g_{c,x},i}, O_{o_\emptyset,g_{c,x},i}\}$ and $\{\Pr(o|g_{c,x}, i)\}$ is a vector of probabilities defined in eqns. (21)-(23).

*Likelihood for outcomes $o_e$ from group $g_{c,x\_u}$*

This grouping aggregates over serostatus $s$ and age $a$, and it focuses on unvaccinated individuals $x_u$ only. The expected hazard rate function for this group is then

$$H_{o_{d,e}, g_{c,x_u}}(t, \boldsymbol{\theta}) = \sum_a p_a \sum_s p_{s,a} H_{o_{d,e}, g_{c,x_u,s,a}}(t, \boldsymbol{\theta}) \quad (25)$$

and

$$H_{o_{h,e}, g_{c,x_u}}(t, \boldsymbol{\theta}) = \sum_a p_a \sum_s p_{s,a} H_{o_{h,e}, g_{c,x_u,s,a}}(t, \boldsymbol{\theta}), \quad (26)$$

where $p_a$ is the proportion of all trial participants in age group $a$ and $p_{s,a}$ is the proportion of trial participants within age group $a$ that have serostatus $s$. Hence, $\Sigma_a p_a = 1$ and $\Sigma_s p_{s,a} = 1$. Due to a lack of information about how the age distribution of trial participants might have varied across countries, we assumed that the overall age distribution reported for the trial as a whole applied to all countries.

Because the outcomes reported are aggregated across non-hospitalized and hospitalized disease, the observed outcomes are $o_1 = o_{d,1} \cup o_{h,1}$, $o_2 = o_{d,2} \cup o_{h,2}$, $o_3 = o_{d,3} \cup o_{h,3}$, $o_4 = o_{d,4} \cup o_{h,4}$, and $o_\varnothing = N_{g,i} - \{o_1 \cup o_2 \cup o_3 \cup o_4\}$. The probabilities corresponding to each of these outcomes are

$$\Pr\left(o_1|g_{c,x},i\right)=\sum_o \Pr\left(o_{o,e}|g_{c,x},i\right),\ (27)$$

$$\Pr\left(o_2|g_{c,x},i\right)=\sum_o \Pr\left(o_{o,e}|g_{c,x},i\right),\ (28)$$

$$\Pr\left(o_3|g_{c,x},i\right)=\sum_o \Pr\left(o_{o,e}|g_{c,x},i\right),\ (29)$$

$$\Pr\left(o_4|g_{c,x},i\right)=\sum_o \Pr\left(o_{o,e}|g_{c,x},i\right),\ (30)$$

and

$$\Pr\left(o_\emptyset|g_{c,x},i\right)=1-\Pr\left(o_1|g_{c,x},i\right)-\Pr\left(o_2|g_{c,x},i\right)-\Pr\left(o_3|g_{c,x},i\right)-\Pr\left(o_4|g_{c,x},i\right).\ (31)$$

The contribution to the likelihood from this portion of the data is then

$$L_{\{o_{d,e}\},g_{c,x_u}}(\boldsymbol{\theta})=\prod_i\prod_s\prod_a Multinomial\left(\{O_{o,g_{c,x_u},i}\}\mid\{\Pr(o|g_{c,x_u},i)\}\right),\ (32)$$

where $\{O_{o,g_{c,x_u},i}\}$ includes only the outcomes $o_1$, $o_2$, $o_3$, $o_4$, and $o_\emptyset = N_{g,i} - o_d$ for group $g_{c,x\_u}$. The probabilities in $\{\Pr(o|g_{c,x\_u},i)\}$ follow eqns. (4) and (5). The only difference is that the hazard rate function from eqns. (25) and (26) are used in the calculation of the survival probabilities in eqns. (6) and (7).

*Likelihood for outcomes $o_{d,e}$, $o_{h,e}$ from group $g_{c\_LKA,x,s}$*

These data aggregate over age groups *a* and are limited to Sri Lanka (ISO3 code LKA). The expected hazard rate functions for this group are then

$$H_{o_{d,e},g_{c_{LKA}},x,s}(t,\boldsymbol{\theta})=\sum_a p_a H_{o_{d,e},g_{c_{LKA}},x,s,a}(t,\boldsymbol{\theta})\ (33)$$

and

$$H_{o_{h,e},g_{c_{LKA}},x,s}(t,\boldsymbol{\theta})=\sum_a p_a H_{o_{h,e},g_{c_{LKA}},x,s,a}(t,\boldsymbol{\theta}),\ (34)$$

where $p_a$ is the proportion of trial participants in age group *a*. Hence, $\Sigma_a\, p_a = 1$. Due to a lack of information about how the age distribution of trial participants might have varied across countries, we assumed that the overall age distribution reported for the trial as a whole applied to all countries.

The contribution to the likelihood from this portion of the data is then

$$L_{\{o_{d,e},o_{h,e}\},g_{c_{LKA}},x,s}(\boldsymbol{\theta})=\prod_i\prod_x\prod_s Multinomial\left(\{O_{o,g_{c_{LKA}},x,s,i}\}\mid\{\Pr(o|g_{c_{LKA}},x,s,i)\}\right),\ (35)$$

where $\{O_{o,g_{x,s},i}\}$ includes all nine outcomes as in eqn. (2) for group $g_{c\_LKA,x,s}$ and $\Pr(o|g_{c\_LKA,x,s},i)$ follows eqns. (4) and (5). The only difference is that the hazard rate functions from eqns. (33) and (34) are used in the calculation of the survival probabilities in eqns. (6) and (7).

*Likelihood for serostatus at baseline s from group $g_c$*

Individuals are seronegative if they have never been exposed to any dengue virus serotype. For a given age group *a*, the probability that an individual is seronegative is simply the product of the probabilities that the individual remains susceptible to each serotype, or $\prod_e \pi_{c,e,a}$. To obtain a probability that applies to all trial participants in country *c*, regardless of age group, we can take the expectation of that probability across the age groups. That results in

$$L_s(\boldsymbol{\theta}) = \prod_c Binomial\left(s_{p,c}, N_c \mid 1 - \sum_a p_a \prod_e \pi_{c,e,\cdot,a}\right), \quad (36)$$

as the contribution to the likelihood from data on serostatus at baseline, where $s_{p,c}$ is the number of $N_c$ total trial participants in country *c* who are seropositive at baseline and $p_a$ is the proportion of all trial participants in age group *a*. Hence, $\Sigma_a\, p_a = 1$. Due to a lack of information about how the age distribution of trial participants might have varied across countries, we assumed that the overall age distribution reported for the trial as a whole applied to all countries.

### 1.3 Derived parameters

Two of the parameters in the hazard rate functions in eqns. (4) and (5) are derived rather than estimated parameters. Below, we explicate the calculation of those derived parameters as a function of estimated parameters.

*Susceptibility*, $\pi_{c,e,s,a}$

Susceptibility, or the probability that a trial participant has not yet been infected with serotype *e*, is a function of the intensity of transmission of that serotype throughout the individual's life and how old the individual is. Because the intensity of DENV transmission can vary over time (36), we considered a force of infection parameter applicable to times prior to the trial, $\bar{\lambda}_{c,e}$, that is distinct from the force of infection during the trial, $\lambda_{c,e}$. This parameter ($\bar{\lambda}_{c,e}$) constitutes the (constant) hazard rate function for infection with serotype *e*. The survival probability of not being infected with serotype *e* prior to age α is then $S(\alpha|c,e,\bar{\lambda}_{c,e}) = \exp(-\bar{\lambda}_{c,e}\alpha)$.

If we momentarily ignore serostatus *s*, we can calculate susceptibility for a randomly selected individual from the population distributed uniformly between ages $a_1$ and $a_2$ as

$$\pi_{c,e,\cdot,a} = \frac{e^{-\bar{\lambda}_{c,e}a_1} - e^{-\bar{\lambda}_{c,e}a_2}}{\bar{\lambda}_{c,e}\left(a_2 - a_1\right)}, \quad (37)$$

which is the expected value of $S(\alpha|c,e,\bar{\lambda}_{c,e})$ across the age range. For seronegative individuals, $\pi_{c,e,s_n,a} = 1$, because they have, by definition, never been infected by any serotype, including *e*. For seropositive individuals, we limit the calculation of susceptibility to individuals who have experienced exactly one prior infection and not two, three, or four prior infections, due to our assumption that post-secondary infections are less likely to result in clinically apparent disease and less likely to be contained within the trial data. We can use eqn. (37) to calculate

$$\pi_{c,e,s_p,a} = \frac{\sum_{f\neq e}\left(\left(1-\pi_{c,f,\cdot,a}\right)\prod_{g\neq f}\pi_{c,g,\cdot,a}\right)}{1-\prod_e \pi_{c,e,\cdot,a}}, \quad (38)$$

which is obtained by taking the probability that an individual has been previously exposed to a serotype other than *e* conditional on being seropositive.

One simplifying assumption of this formulation is that it slightly overrepresents $\pi_{c,e,\cdot,a}$ for a given value of $\bar{\lambda}_{c,e}$, because it ignores the fact that some portion of individuals will be temporarily cross-immune to serotype *e* due to recent infection with some other serotype. In practice, the proportion who are temporarily cross-immune at a given time is likely to be relatively small, although it could lead to some slight overestimation of $\bar{\lambda}_{c,e}$ relative to its true value if temporary cross-immunity were accounted for. Given that estimation of $\bar{\lambda}_{c,e}$ is not of primary interest, we viewed this as an acceptable compromise for the sake of simplifying the calculations.

*Relative risk*, $RR_{o,x}$

The initial level of protection afforded by vaccination is defined as the initial relative risk, ϕ. As time passes, relative risk approaches 1 due to waning at rate δ, which results in

$$RR_{o_d,x_v}(t, \phi_{o_d,s,e}, \delta_{o_d,s}) = 1 - \phi_{o_d,s,e} e^{-\delta_{o_d,s} t} \quad (39)$$

for relative risk of non-hospitalized disease and

$$RR_{o_h,x_v}(t, \phi_{o_h,s,e}, \delta_{o_h,s}) = 1 - \phi_{o_h,s,e} e^{-\delta_{o_h,s} t} \quad (40)$$

for relative risk of hospitalized disease, both of which apply to vaccinated individuals only. As the notation in eqns. (39) and (40) indicates, the initial relative risk depends on serostatus and serotype, and the rate of waning depends on serostatus. For unvaccinated individuals,

$$RR_{o_d,x_u} = RR_{o_h,x_u} = 1 \quad (41)$$

because there is no vaccine and, therefore, no modification of risk.

We can also denote time-varying per-exposure protection against non-hospitalized disease as

$$PEP_{o_d,x_v}(t, \phi_{o_d,s,e}, \delta_{o_d,s}) = \phi_{o_d,s,e} e^{-\delta_{o_d,s} t}$$

or

$$PEP_{o_d,x_v}(t, \phi_{o_d,s,e}, \delta_{o_d,s}) = 1 - RR_{o_d,x_v}(t, \phi_{o_d,s,e}, \delta_{o_d,s}) .$$

Similarly, we can denote time-varying per-exposure protection against hospitalized disease as

$$PEP_{o_h,x_v}(t, \phi_{o_h,s,e}, \delta_{o_h,s}) = \phi_{o_h,s,e} e^{-\delta_{o_h,s} t}$$

or

$$PEP_{o_h,x_v}(t, \phi_{o_h,s,e}, \delta_{o_h,s}) = 1 - RR_{o_h,x_v}(t, \phi_{o_h,s,e}, \delta_{o_h,s}) .$$

The above equations inform only on the relative risk at a given instant *t*. Because the intervals over which the data are censored are potentially somewhat long relative to the timescale of waning, we must account for the time-varying nature of relative risk in the likelihood. This occurs in eqn. (6) through calculation of the cumulative hazard function. Since $\int H_{o,g}\,dt$ does not have a closed form solution, we instead approximated the cumulative hazard function using a Riemann sum, such that

$$\int_0^t H_{o,g}(t, \boldsymbol{\theta})dt \approx \sum_{i=1}^{n} H_{o,g}(t_i, \boldsymbol{\theta})\Delta t_i \quad (42)$$

where $\Delta t_i$ is one month, or 1/12.

### 1.4 Independent-parameter model

For the independent-parameter model, we only incorporated serotype- and serostatus-specific data; i.e. data on outcomes $o_{o,e}$ from group $g_{x,s}$, aggregated across time. To minimize parameters and reduce complexity of the model, rather than calculating a hazard function and then deriving a survival probability based on that survival function, we directly estimated the baseline probability that an individual in the placebo arm with serostatus *s* experience an outcome *o* due to infection with serotype *e*, which we denote as $\Pr(o_{o,e}|g_{x\_u,s})$. To align with assumptions made when reported efficacy estimates are calculated, we assumed that these probabilities are independent of one another, thereby minimizing the degree to which the data could jointly inform those parameters.

The only way in which data were allowed to jointly inform the parameters in the independent-parameter model was via our assumption that the probability that an individual in the vaccine arm with serostatus *s* experiences an outcome *o* due to infection with serotype *e* is a function of the baseline probability and an efficacy parameter $\phi_{o,s,e}$, such that

$$Pr(o_{o,e}|g_{x_v,s}) = (1 - \phi_{o,s,e})Pr(o_{o,e}|g_{x_a,s}) \quad (43).$$

The assumption of a shared baseline probability aligns with the assumptions of Cox proportional hazards analysis, which was used to calculate efficacy in the trial.

Our likelihood is then

$$L_{\{o_{d,e}, o_{h,e}\}, g_{x,s}}(\boldsymbol{\theta}) = \prod_x \prod_s Multinomial\left(\{O_{o,g_{x,s},i}\} \mid \{\Pr(o|g_{x,s})\}\right), \quad (44)$$

where $\{O_{o,g_{x,s},i}\}$ includes all nine outcomes as in eqn. (2) for group $g_{x,s}$ and $\Pr(o|g_{x\_v,s})$ follows eqn. (43).

### 2. Supplemental Results

2.1 Estimates of per-exposure protection

***Table S4.*** *Per-exposure protection against non-hospitalized disease at initial vaccination and ten years post-vaccination.*

| | Seronegative | | Seropositive | |
|---|---|---|---|---|
| | Median at initial vaccination (95% CrI) | Median at ten years post-vaccination (95% PPI) | Median at initial vaccination (95% CrI) | Median at ten years post-vaccination (95% PPI) |
| DENV-1 | 49.8 (31.0, 68.1) | 21.5 (12.3, 30.6) | 75.2 (64.0, 86.4) | 28.1 (21.2, 35.1) |
| DENV-2 | 99.6 (96.4, 100.0) | 42.6 (26.1, 57.6) | 99.5 (96.3, 100.0) | 35.7 (24.6, 46.9) |
| DENV-3 | 26.7 (-8.2, 53.4) | 11.0 (-3.5, 24.8) | 46.1 (31.0, 61.1) | 16.3 (9.5, 23.6) |
| DENV-4 | 38.1 (-28.6, 100.0) | 15.7 (-16.7, 38.5) | 83.9 (57.4, 100.0) | 29.4 (17.6, 41.6) |

***Table S5.*** *Per-exposure protection against hospitalized disease at initial vaccination and ten years post-vaccination.*

| | Seronegative | | Seropositive | |
|---|---|---|---|---|
| | Median at initial vaccination (95% CrI) | Median at ten years post-vaccination (95% PPI) | Median at initial vaccination (95% CrI) | Median at ten years post-vaccination (95% PPI) |
| DENV-1 | 90.6 (65.5, 100.0) | 31.7 (14.5, 49.1) | 49.9 (17.3, 82.0) | 17.9 (5.1, 31.1) |
| DENV-2 | 98.0 (83.0, 100.0) | 34.9 (15.8, 54.4) | 98.8 (90.1, 100.0) | 35.0 (16.9, 52.5) |
| DENV-3 | -72.7 (-149.4, -4.0) | -25.4 (-59.9, -1.1) | 52.3 (10.9, 88.4) | 17.8 (2.9, 34.0) |
| DENV-4 | 90.8 (28.4, 100.0) | 30.4 (5.2, 53.5) | 90.3 (22.0, 100.0) | 29.6 (3.8, 52.0) |

2.2 Model predictions compared to trial data by data type

*Non-hospitalized and hospitalized disease by serotype stratified by vaccination status and serostatus*

The number of non-hospitalized and hospitalized cases in each arm of the trial by infecting serotype and baseline serostatus was available from the fourth month (one month after the second dose) to the 57th month in the trial (Fig. S2). The model was fit to the data from each time interval available, which were months 4 to 16, months 16 to 27, months 27 to 39, months 39 to 50, and months 50 to 57 (Figs. S11-S14). Here, we present model predictions of cumulative cases across all available time intervals and data. Case counts were generally higher in seropositive individuals across arms. DENV-2 and DENV-1 caused the most cases, whereas DENV-4 caused the fewest. Overall, the model generally captures the patterns in the data, although the model slightly overpredicts hospitalizations due to DENV-2.

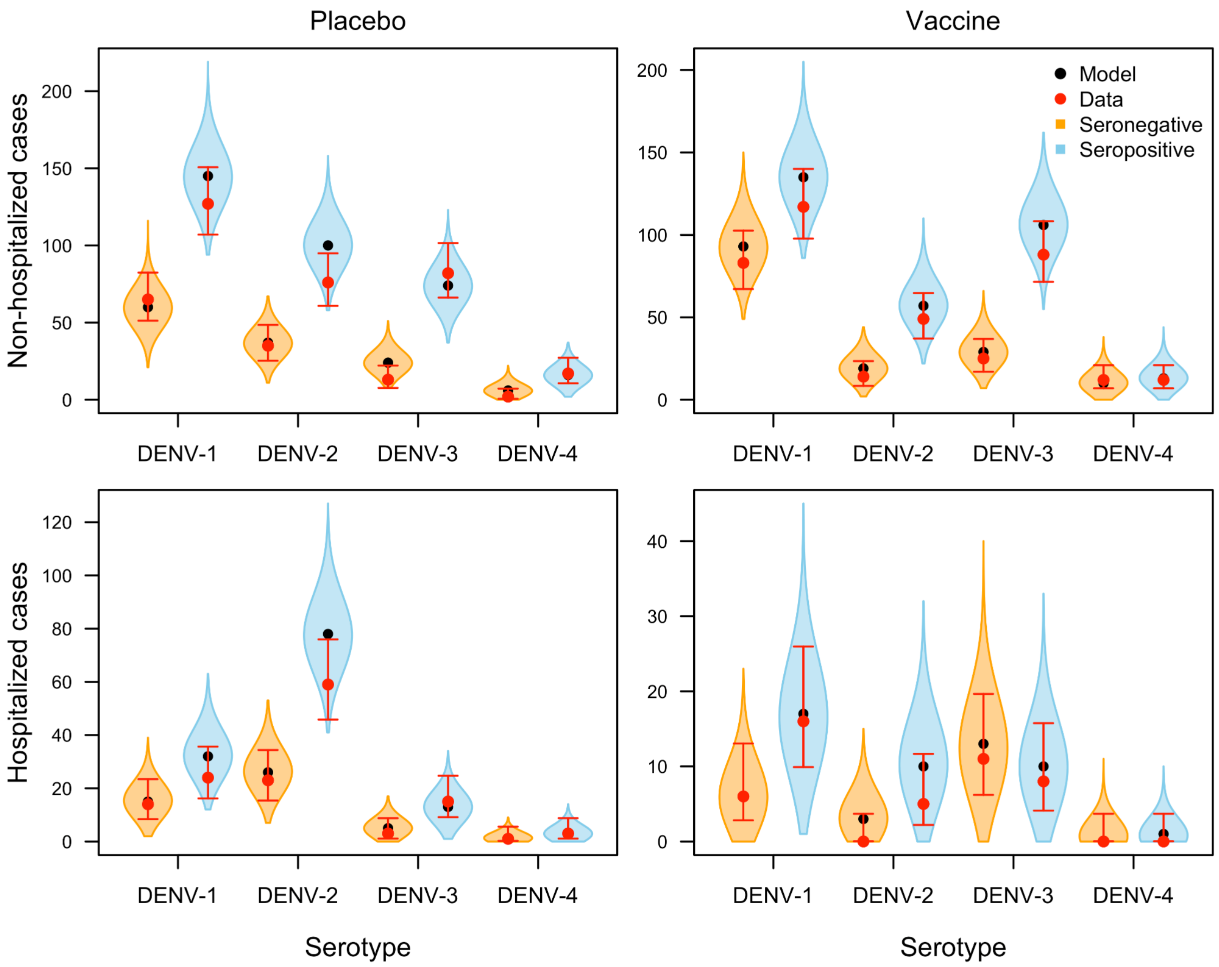


***Figure S2.*** *Posterior predictions of the number of non-hospitalized (top row) and hospitalized (bottom row) cases by serotype (x-axis) and serostatus (colors) from 4 to 57 months after the first vaccine dose in the placebo (left column) and vaccine (right column) arms. Violin shapes show how the model's posterior predictions were distributed, with black dots indicating median predictions. The red dot and lines show the observed value in the trial and the 95% posterior*

*predictive interval around that observation, which was calculated assuming a beta conjugate prior (α=1, β=1) and a binomial likelihood to account for the sampling process and associated uncertainty involved in collecting the data.*

*Non-hospitalized and hospitalized disease stratified by vaccination status, serostatus, and age*
Data on the number of cases by age and serostatus was available from the fourth month (one month after second dose) to 57th month in the trial (Fig. S3). Individuals in the trial were broken down into three age groups: 4-5 years old, 6-11 years old, and 12-16 years old. The model was fit to the data from each time interval available, which were months 4 to 16, months 16 to 27, months 27 to 39, months 39 to 50, and months 50 to 57 (Figs. S15-S18). Here, we present model predictions of cumulative cases across all available time intervals and data. The number of cases was highest in the 6-11 year old age group (also the largest age group in terms of number of participants). Model predictions generally align with the patterns in the data, given that there were no age-specific parameters directly estimated by the model.

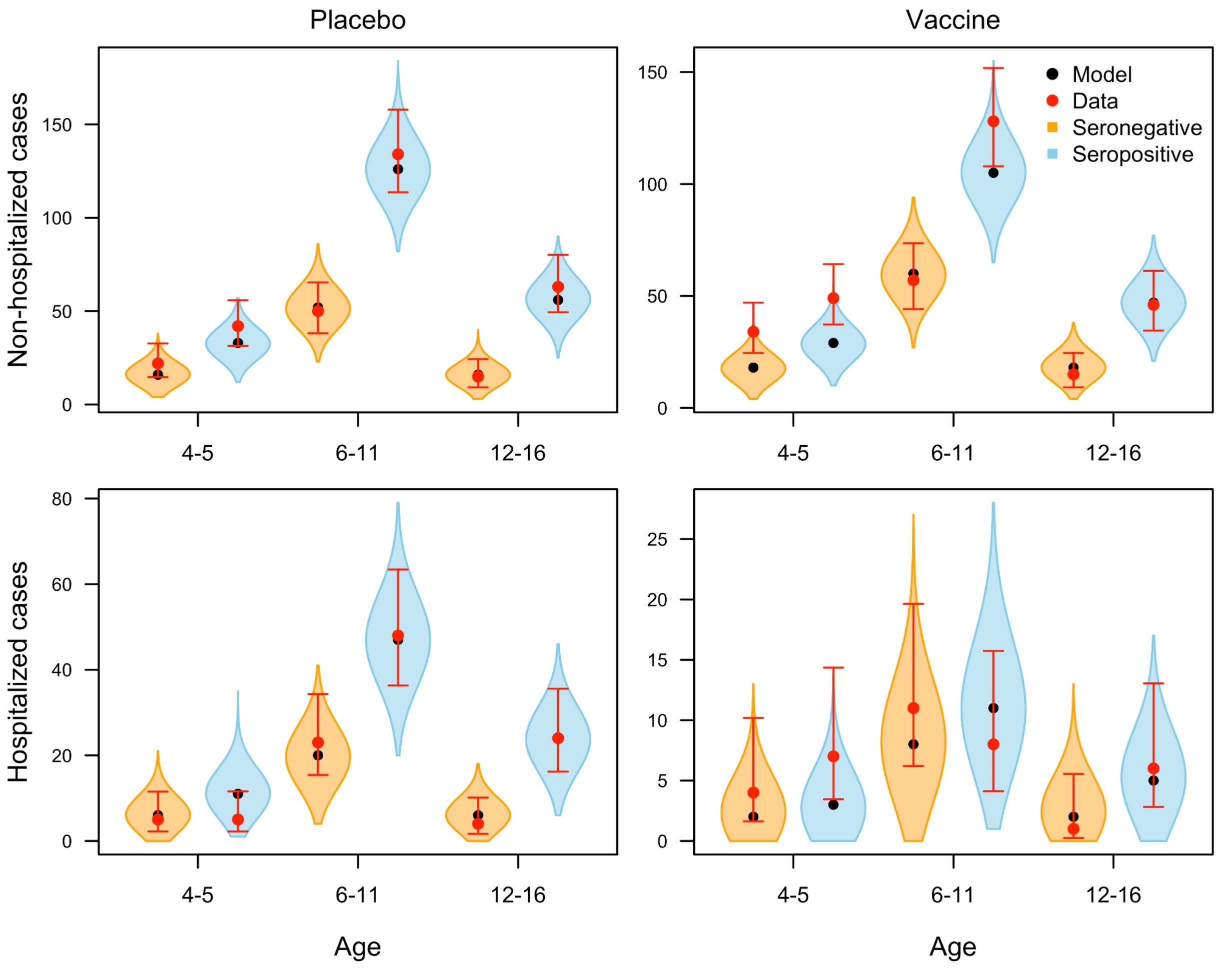


***Figure S3.*** *Posterior predictions of the number of non-hospitalized (top row) and hospitalized (bottom row) cases in each age group in the trial (x-axis) by serostatus (colors) from 4 to 57 months after the first vaccine dose in the placebo (left column) and vaccine (right column) arms.*

*Violin shapes show how the model's posterior predictions were distributed, with black dots indicating median predictions. The red dot and lines show the observed value in the trial and the 95% posterior predictive interval around that observation, which was calculated assuming a beta conjugate prior (α=1, β=1) and a binomial likelihood to account for the sampling process and associated uncertainty involved in collecting the data.*

*Non-hospitalized and hospitalized disease stratified by country and vaccination status*
The numbers of non-hospitalized cases in the placebo arm by country were available for the first 39 months of the trial, starting with the administration of the first dose. Here, we present model predictions of cumulative cases across all available time intervals and data. Overall, the Philippines had the most cases, while Nicaragua had the fewest. The heterogeneity by country was captured by the model predictions (Fig. S4). Hospitalizations were highest in Sri Lanka among all eight countries in the trial. The model predictions capture the trends in the data.

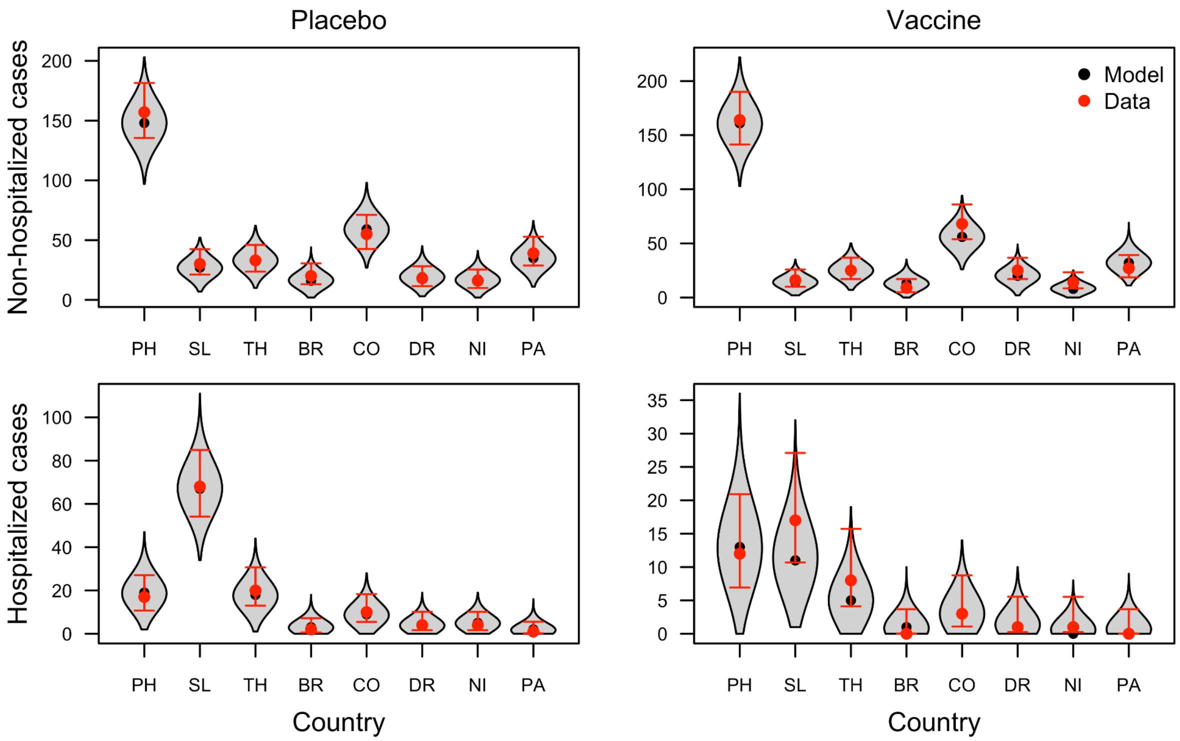


***Figure S4.*** *Posterior predictions of the number of non-hospitalized (top row) and hospitalized (bottom row) cases in each country in the trial (Philippines, Sri Lanka, Thailand, Brazil, Colombia, Dominican Republic, Nicaragua, and Panama) from 1 to 39 months after the first vaccine dose in the placebo (left column) and vaccine (right column) arms. Violin shapes show how the model's posterior predictions were distributed, with black dots indicating median predictions. The red dot and lines show the observed value in the trial and the 95% posterior predictive interval around that observation, which was calculated assuming a beta conjugate prior (α=1, β=1) and a binomial likelihood to account for the sampling process and associated uncertainty involved in collecting the data.*

*Disease by serotype stratified by country for unvaccinated individuals*
Serotype-specific case data were available by country, although these data were only available aggregated across non-hospitalized and hospitalized cases, for the placebo arm only, and for the fourth month (one month after the second dose) to 39th month in the trial. In the Philippines, most cases were due to DENV-3. In both Sri Lanka and Nicaragua, DENV-2 was the dominant serotype. In Thailand and Brazil, most cases were due to either DENV-1 or DENV-2. For the remaining countries (Colombia, Dominican Republic, and Panama), most cases were due to DENV-1.

The model fit well to most data, but slightly underpredicted cases for each of the dominant serotypes in each country, particularly DENV-3 in the Philippines and DENV-2 in Sri Lanka (Fig. S5). Given that these data were relatively coarse, as cases were aggregated by outcomes and only available for a single time point, these data may have not influenced the likelihood calculation as much as other data types. Therefore, the model may have been influenced more by other data types.

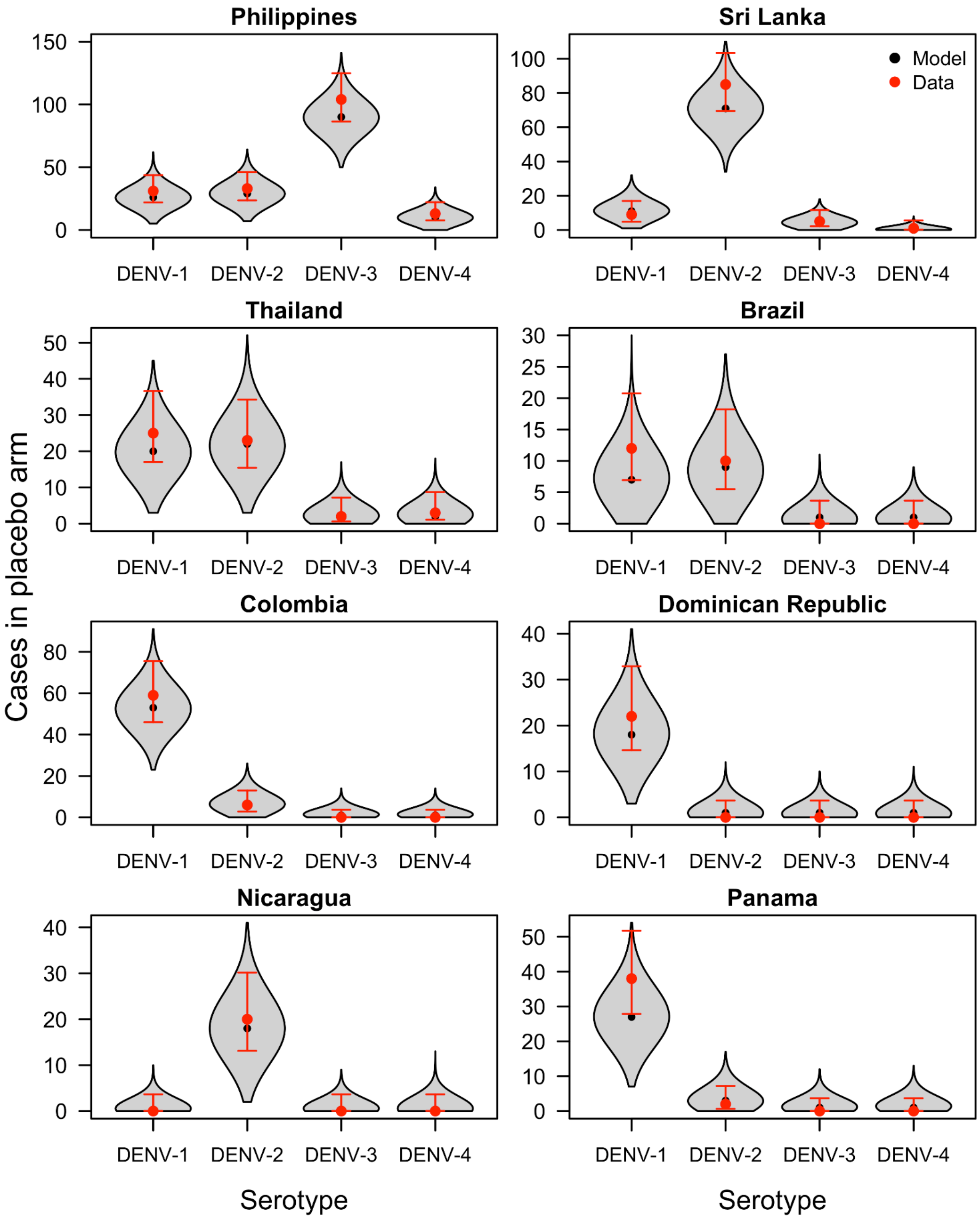


***Figure S5.*** *Posterior predictions of the number of cases in each country in the trial (panels) by serotype (x-axis) from 4 to 39 months after the first vaccine dose in the placebo arm. Violin shapes show how the model's posterior predictions were distributed, with black dots indicating median predictions. The red dot and lines show the observed value in the trial and the 95% posterior predictive interval around that observation, which was calculated assuming a beta conjugate prior (α=1, β=1) and a binomial likelihood to account for the sampling process and associated uncertainty involved in collecting the data.*

*Non-hospitalized and hospitalized disease by serotype for Sri Lanka stratified by serostatus and vaccination status*

Data were available on non-hospitalized and hospitalized cases in the vaccine and placebo arms by infecting serotype and baseline serostatus for Sri Lanka for the fourth month (one month after the second dose) to 57th month in the trial (Fig. S6). DENV-2 remained the dominant serotype from the 39th to 57th month in the trial (Fig. S6). Overall, the model captures most of the data, but slightly overestimates cases in the vaccine arm. The model may have estimated a relatively high trial force of infection of DENV-2 in Sri Lanka to capture the relatively high case counts, particularly in the placebo arm, but based on other data types (such as data by baseline serostatus and infecting serotype), did not estimate a high enough protection against DENV-2 to reproduce sufficiently low DENV-2 cases in Sri Lanka specifically.

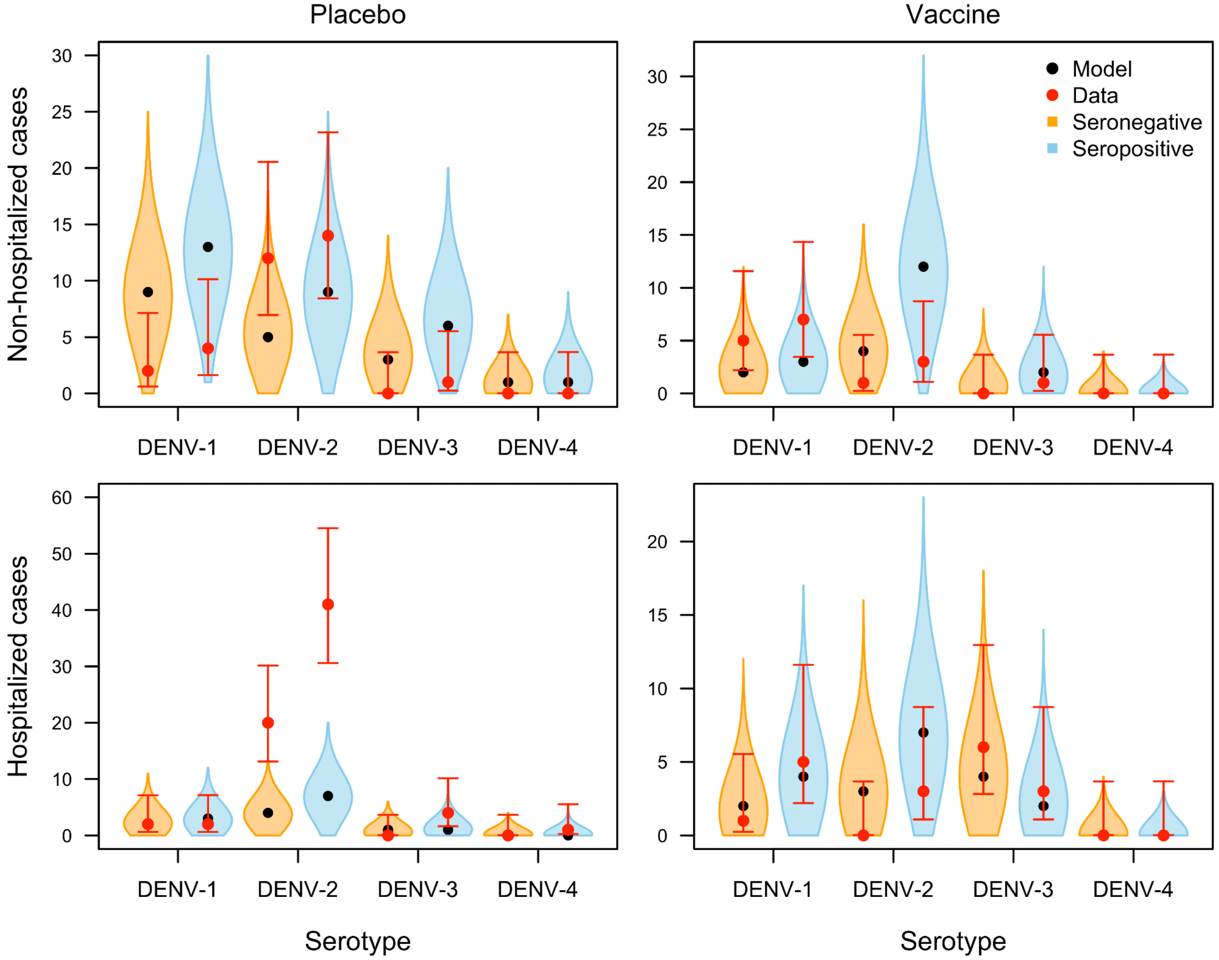


***Figure S6.*** *Posterior predictions of the number of non-hospitalized (top row) and hospitalized (bottom row) cases in Sri Lanka by serotype (x-axis) and serostatus (color) from 4 to 57 months after the first vaccine dose in the placebo (left column) and vaccine (right column) arms. Violin shapes show how the model's posterior predictions were distributed, with black dots indicating median predictions. The red dot and lines show the observed value in the trial and the 95% posterior predictive interval around that observation, which was calculated assuming a beta*

*conjugate prior (α=1, β=1) and a binomial likelihood to account for the sampling process and associated uncertainty involved in collecting the data.*

*Serostatus at baseline stratified by country*

Data were available on the number of participants by country who were seropositive at baseline, which was defined as having a reciprocal neutralizing antibody titer ≥10). The proportion of participants in each country who were seropositive was highest in the Dominican Republic, with 97.2% of participants being seropositive at baseline, followed by the Philippines (87.6%), Colombia (84.6%), Nicaragua (77.7%), Brazil (71.2%), Thailand (65.6%), Sri Lanka (61.5%), and Panama (37.8%). The model captures this data (Fig. S7).

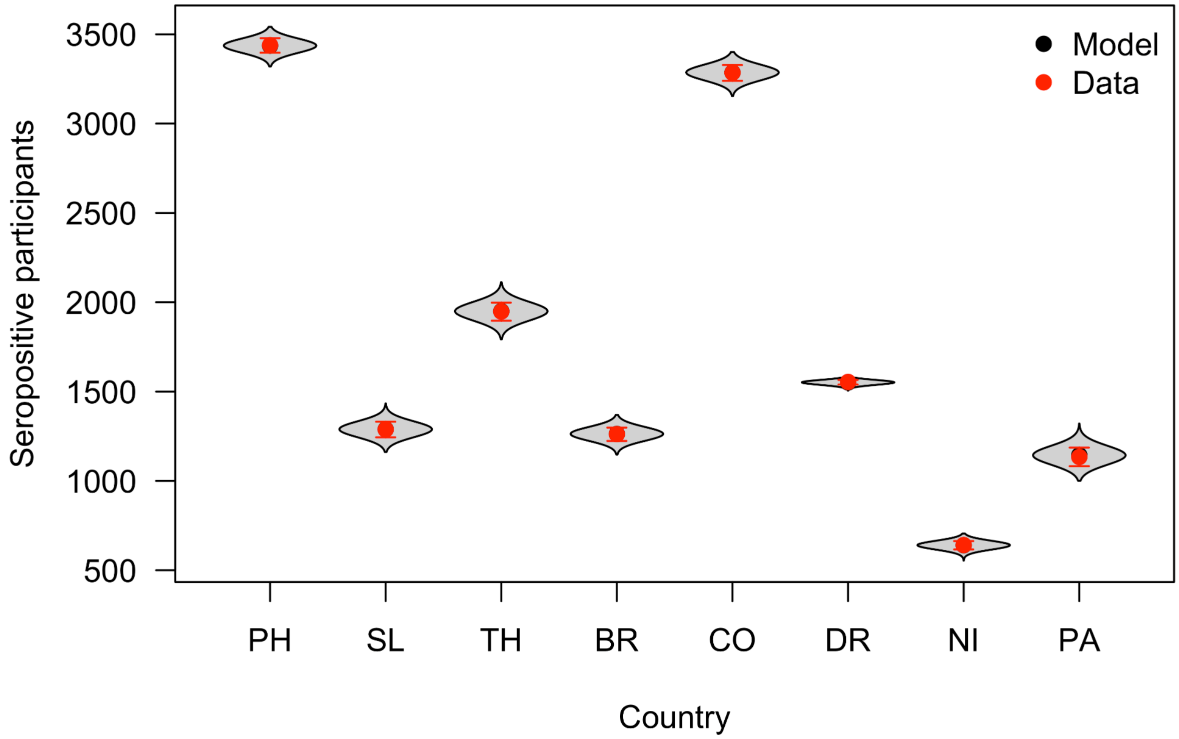


***Figure S7.*** *Posterior predictions of the number of seropositive participants (y-axis) in each country in the trial (x-axis) at baseline. Violin shapes show how the model's posterior predictions were distributed, with black dots indicating median predictions. The red dot and lines show the observed value in the trial and the 95% posterior predictive interval around that observation, which was calculated assuming a beta conjugate prior (α=1, β=1) and a binomial likelihood to account for the sampling process and associated uncertainty involved in collecting the data.*

### 2.3 Epidemiological parameter estimates

*Historical force of infection*

We estimated the background force of infection prior to the trial by each country and serotype to allow us to account for pre-existing serotype-specific immunity (Fig. S8). Overall, the model estimated the highest annual forces of infection for the Dominican Republic (DENV-1: 0.05 [95%

CrI: 0.01, 0.17], DENV-2: 0.12 [95% CrI: 0.01, 0.31], DENV-3: 0.10 [95% CrI: 0.01, 0.29], DENV-4: 0.11 [95% CrI: 0.01, 0.30), which aligns with the fact that the Dominican Republic had the highest proportion of seropositive individuals of the eight countries in the trial. Generally, there was little heterogeneity in serotype-specific historical force of infection by country, which could partially be attributed to the fact that data on baseline serostatus was not serotype-specific. One exception is the Philippines, in which the historical force of infection for DENV-3 (0.01, 95% CrI: [0.004, 0.02]) is relatively low compared to the other three serotypes; this aligns with the fact that cases due to DENV-3 were relatively high in the Philippines, thereby indicating that pre-existing DENV-3-specific immunity should be relatively low among participants.

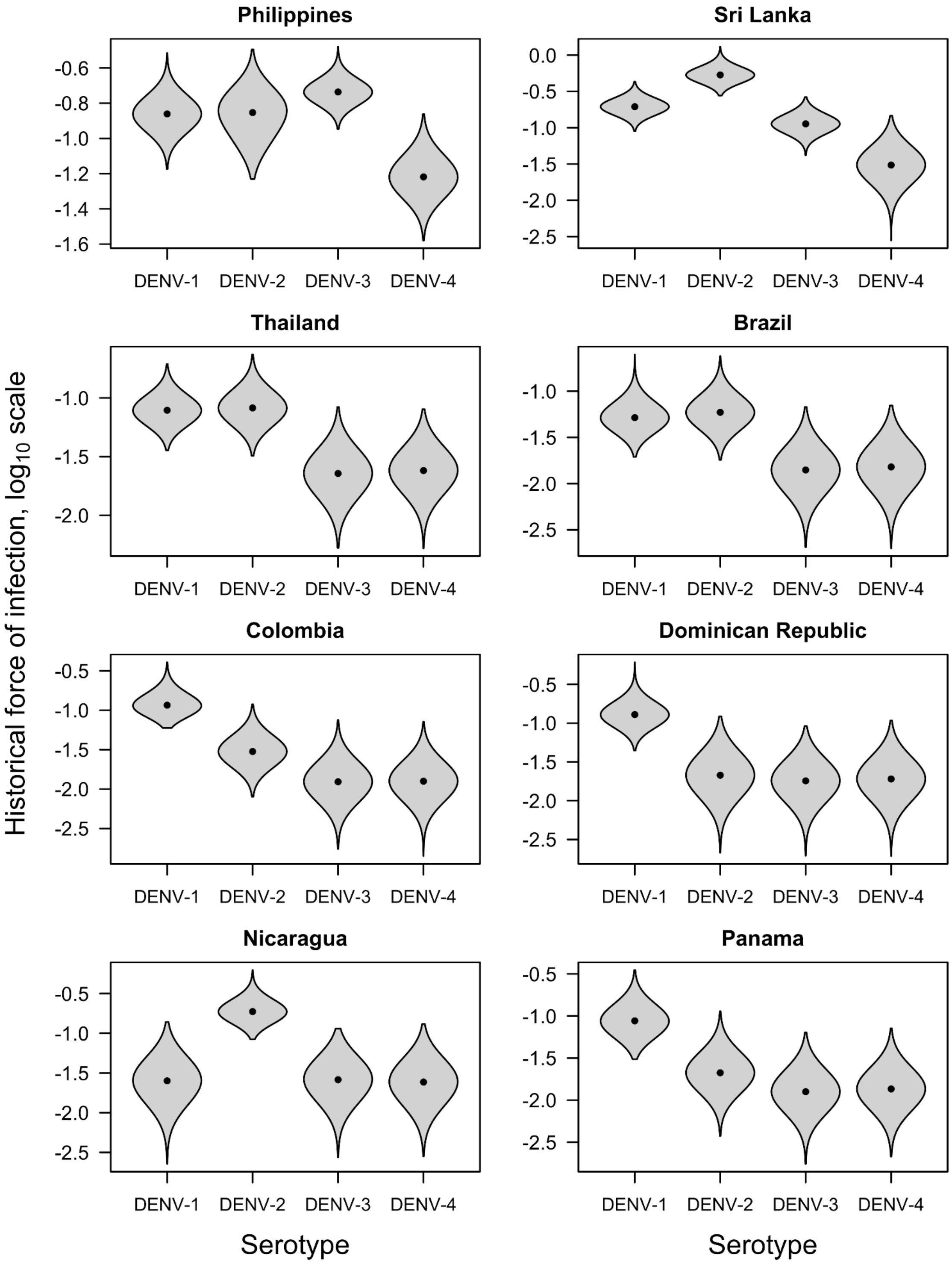


***Figure S8.*** *Force of infection prior to the trial by serotype (x-axis) and by each country in the trial (panels) presented on a* $log_{10}$ *scale. Violin shapes show how the model's posterior estimates were distributed, with black dots indicating median estimates.*

*Force of infection during the trial*

We estimated the force of infection during the trial by serotype and by country (Figure S9). As expected, within each country, serotypes for which higher forces of infection were estimated aligned with serotypes that resulted in more cases (Fig. S9). For example, cases due to DENV-2 were most prevalent in Sri Lanka, and the estimated trial force of infection of DENV-2 in Sri Lanka was 0.53 (95% CrI: 0.36, 0.75). Similarly, DENV-3 was the dominant serotype in the Philippines, and the estimated trial force of infection of DENV-3 in the Philippines was estimated to be 0.18 (95% CrI: 0.14, 0.23).

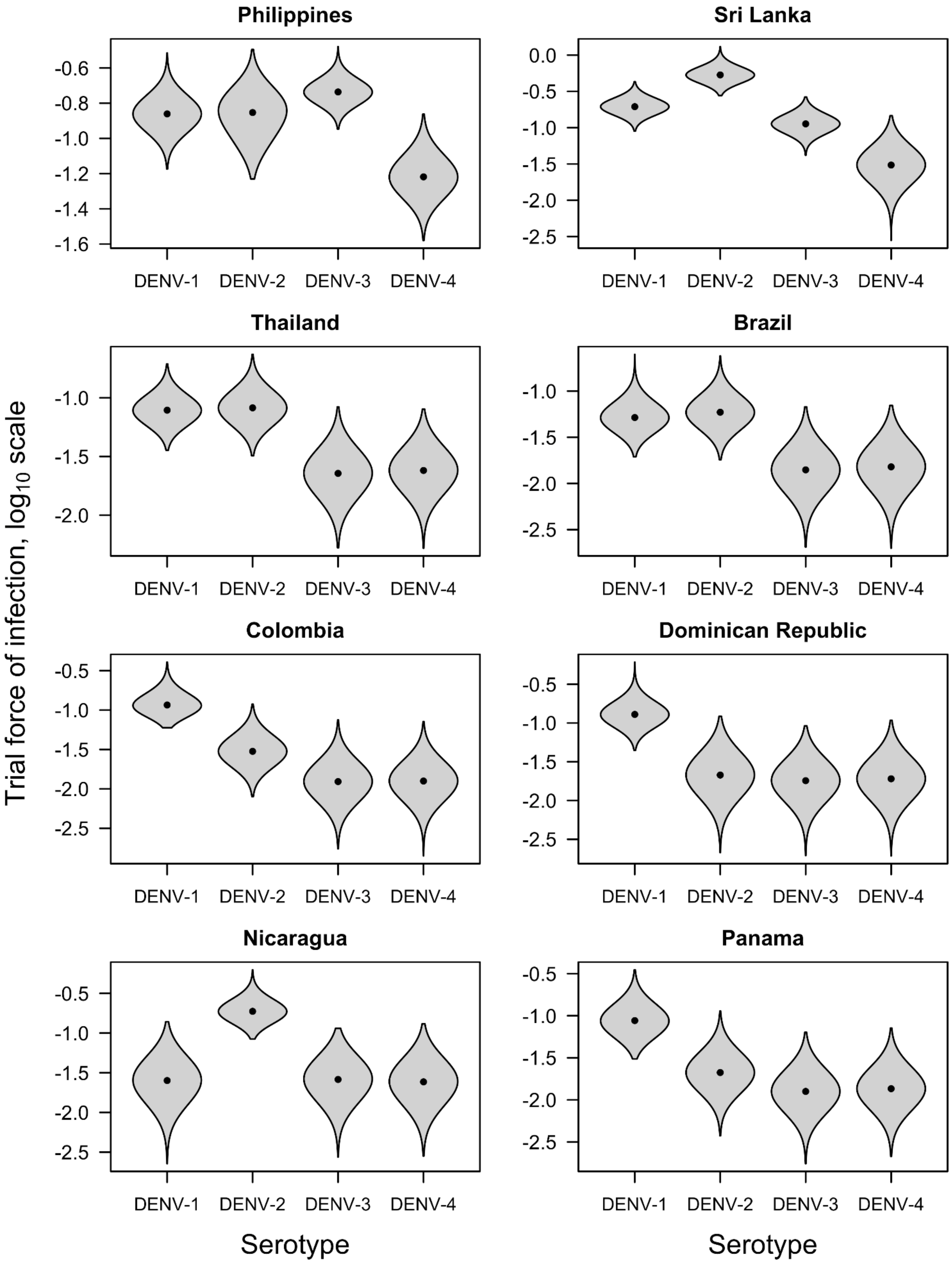


***Figure S9.*** *Force of infection during the trial by serotype (x-axis) and by each country in the trial (panels) presented on a $log_{10}$ scale. Violin shapes show how the model's posterior estimates were distributed, with black dots indicating median estimates.*

*Baseline probability of outcomes by infection valency*

We estimated the probability of non-hospitalized disease and hospitalized disease for primary and secondary infections (Fig. S10). The probability of non-hospitalized disease given infection is higher for individuals experiencing secondary infections (0.83, 95% CrI: 0.73, 0.92) than individuals experiencing primary infections (0.33, 95% CrI: 0.27, 0.39). This aligns with the general understanding that secondary dengue infections are generally more likely to cause symptomatic disease due to antibody-dependent enhancement (29).

The probability of hospitalized disease given disease is similar for individuals experiencing primary infections (0.29, 95% CrI: 0.21, 0.38) and individuals experiencing secondary infections (0.24, 95% CrI: 0.18, 0.31). This may be due to the fact that hospitalization may be affected by confounding factors that impact admission, rather than infection valency alone (12,32).

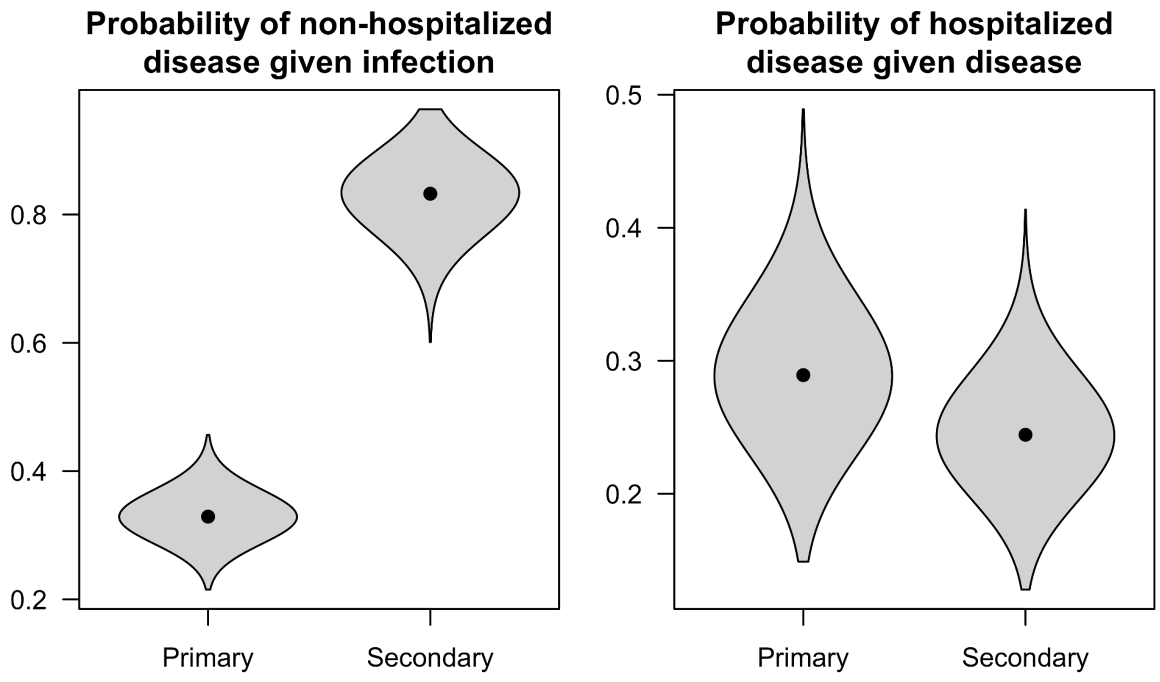


***Figure S10.*** *Baseline probability of non-hospitalized disease given infection (left) and probability of hospitalized disease given disease (right) by infection valency (x-axis). Violin shapes show how the model's posterior estimates were distributed, with black dots indicating median estimates.*

*Country-specific reporting rates of outcomes by country*

We estimated a country-specific probability of reporting each outcome, disease and severe disease to account for country-specific differences in surveillance and clinical care (Fig. S10). In the context of non-hospitalized disease, the reporting probability refers to the probability that a non-hospitalized case is accurately detected and reported. On the other hand, in the context of severe disease, the reporting probability refers to the probability that a severe case is hospitalized and thereby recorded as a hospitalization. We find that most countries have similarly high probabilities of reporting non-hospitalized disease, with the exception of Sri Lanka (0.13, 95% CrI: 0.07, 0.19). On the other hand, Sri Lanka had the highest rate of reporting severe disease (0.97, 95% CrI: 0.86, 0.99). Probabilities of reporting hospitalized disease are estimated to be lower than probabilities of reporting non-hospitalized disease on average, which aligns with our expectation that non-hospitalized cases were better recorded due to active surveillance during the trial, whereas hospitalization rates were dependent on local practices (16).

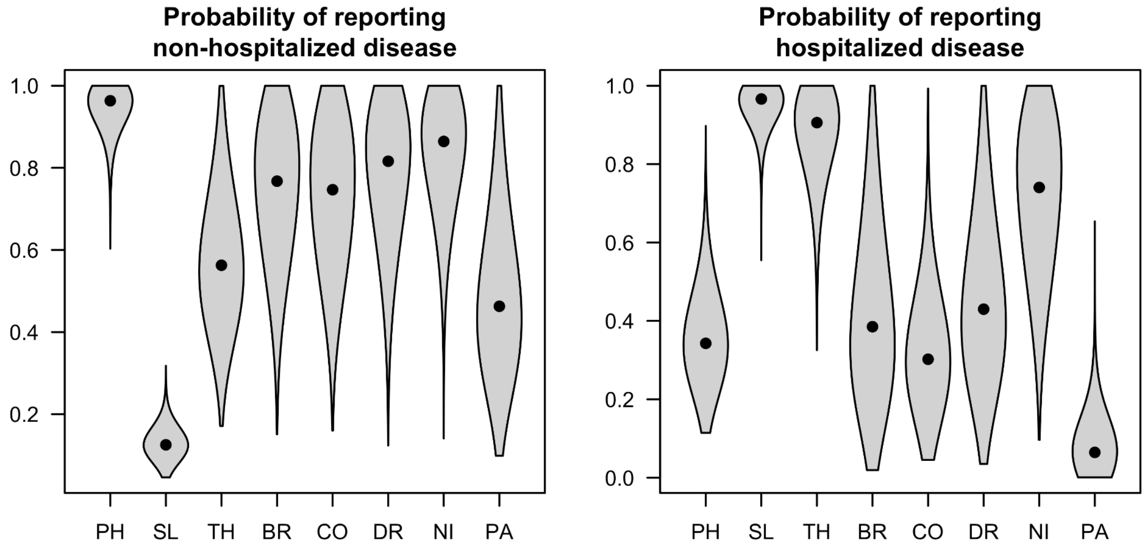


***Figure S11.*** *Probability of reporting non-hospitalized disease (left) and hospitalized disease (right) by each of the eight countries in the trial (x-axis). Violin shapes show how the model's posterior estimates were distributed, with black dots indicating median estimates.*

### 2.4 Model predictions broken down by time intervals

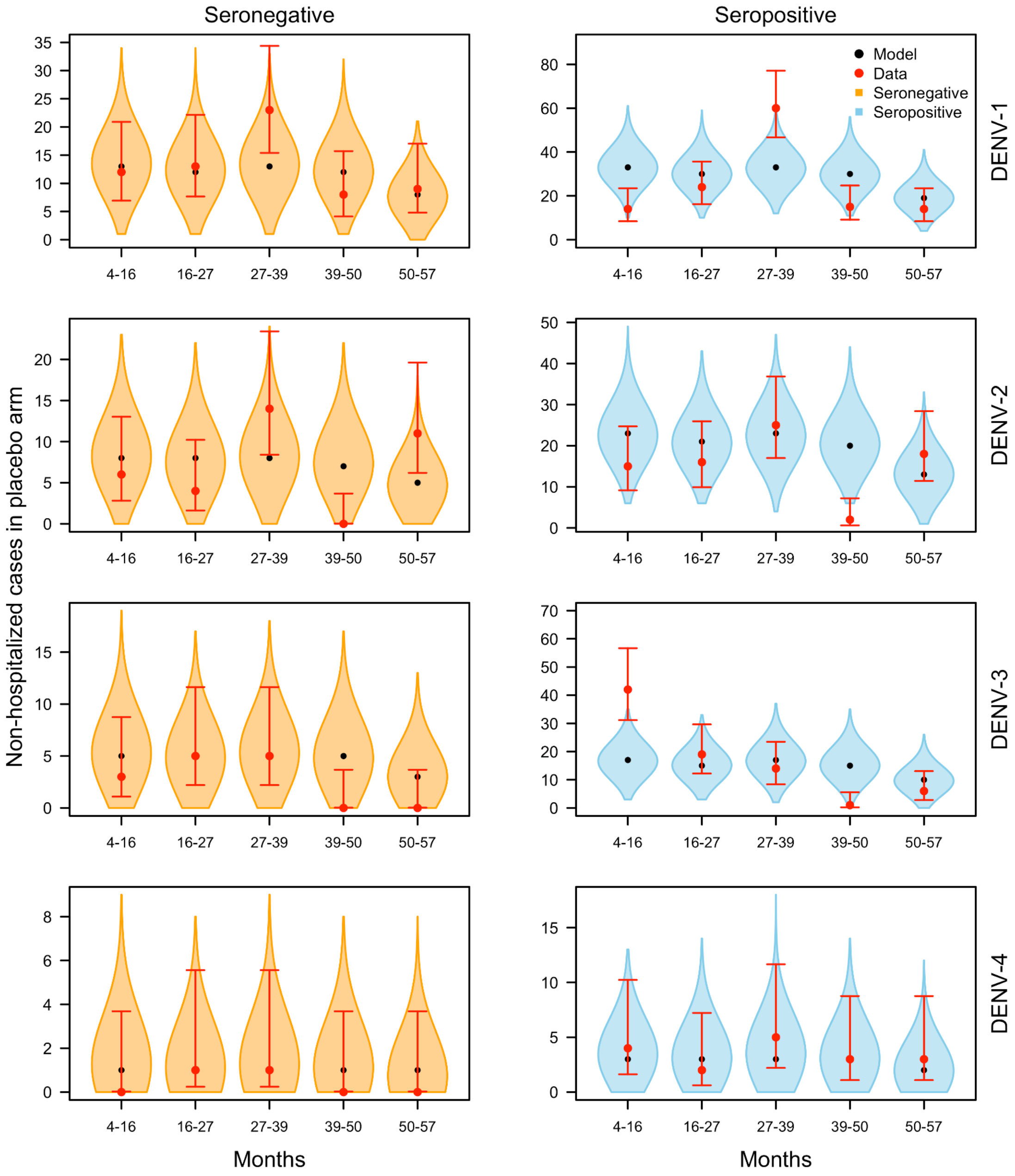


***Figure S12.*** *Posterior predictions of the number of non-hospitalized cases by serotype (rows) and serostatus (columns) from 4 to 57 months (x-axis) after the first vaccine dose in the placebo arm over time. Violin shapes show how the model's posterior predictions were distributed, with black dots indicating median predictions. The red dot and lines show the observed value in the trial and the 95% posterior predictive interval around that observation, which was calculated assuming a beta conjugate prior (α=1, β=1) and a binomial likelihood to account for the sampling process and associated uncertainty involved in collecting the data.*

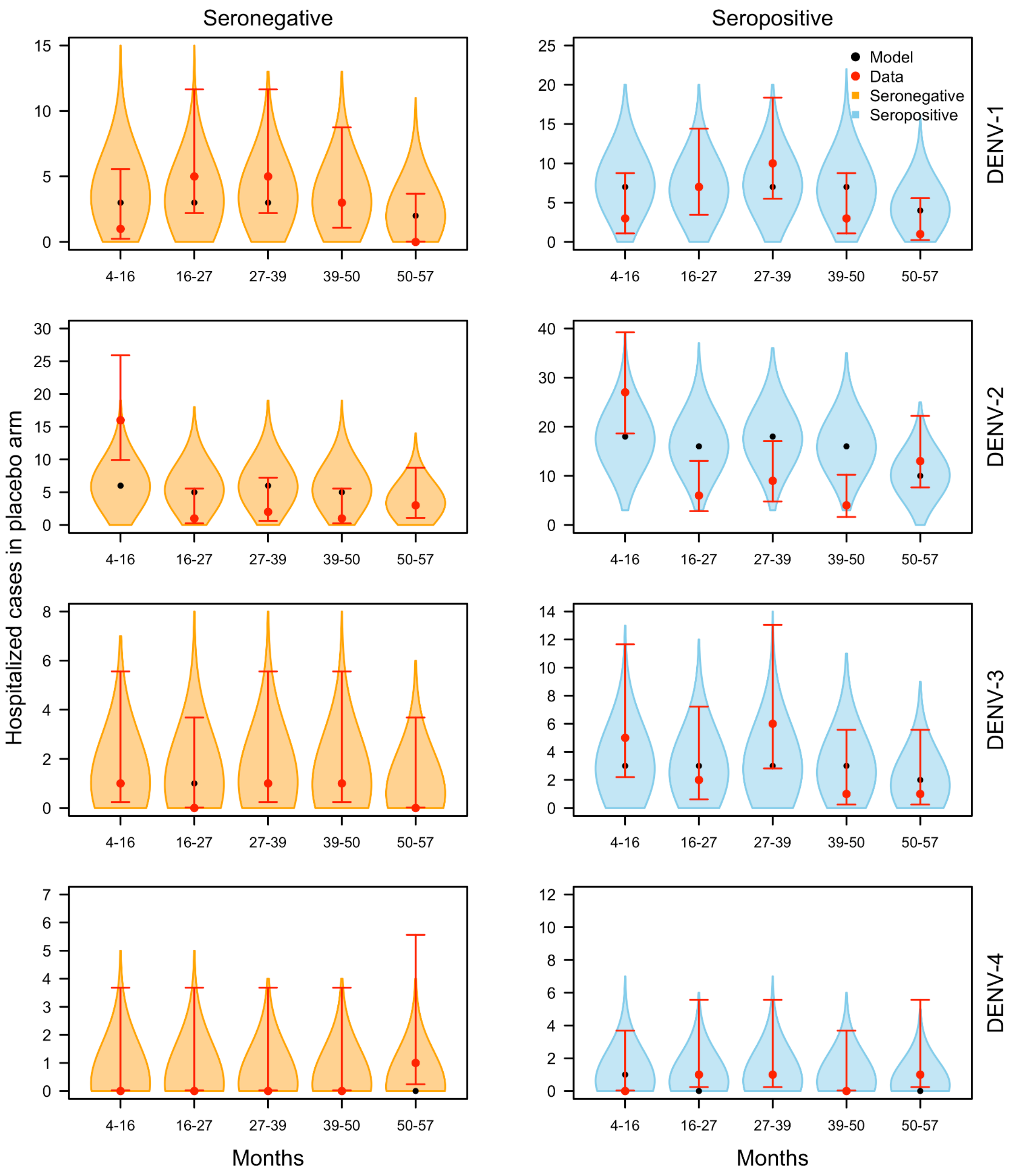


***Figure S13.*** *Posterior predictions of the number of hospitalized cases by serotype (rows) and serostatus (columns) from 4 to 57 months (x-axis) after the first vaccine dose in the placebo arm over time. Violin shapes show how the model's posterior predictions were distributed, with black dots indicating median predictions. The red dot and lines show the observed value in the trial and the 95% posterior predictive interval around that observation, which was calculated assuming a beta conjugate prior (α=1, β=1) and a binomial likelihood to account for the sampling process and associated uncertainty involved in collecting the data.*

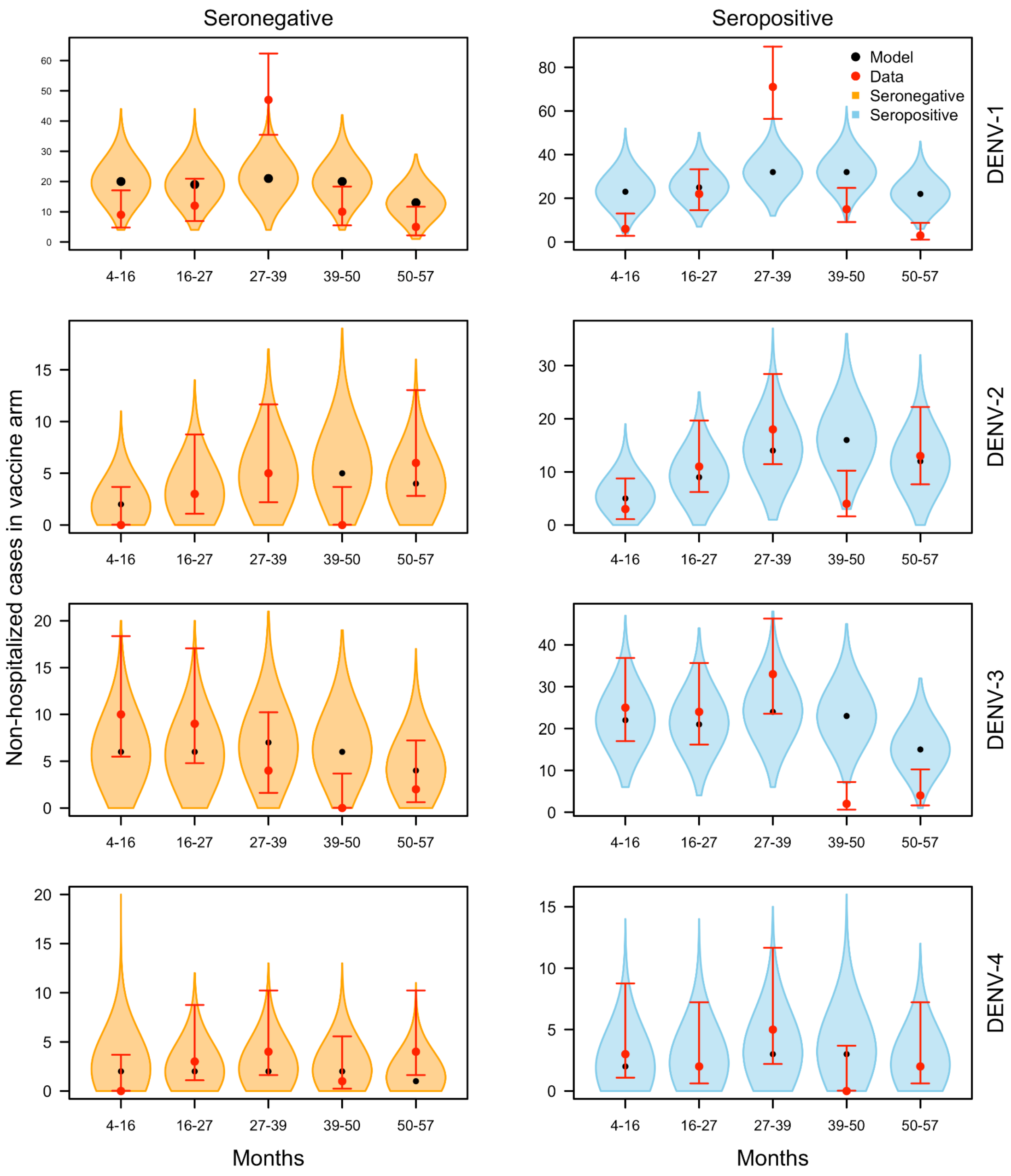


***Figure S14.*** *Posterior predictions of the number of non-hospitalized cases by serotype (rows) and serostatus (columns) from 4 to 57 months (x-axis) after the first vaccine dose in the vaccine arm over time. Violin shapes show how the model's posterior predictions were distributed, with black dots indicating median predictions. The red dot and lines show the observed value in the trial and the 95% posterior predictive interval around that observation, which was calculated assuming a beta conjugate prior (α=1, β=1) and a binomial likelihood to account for the sampling process and associated uncertainty involved in collecting the data.*

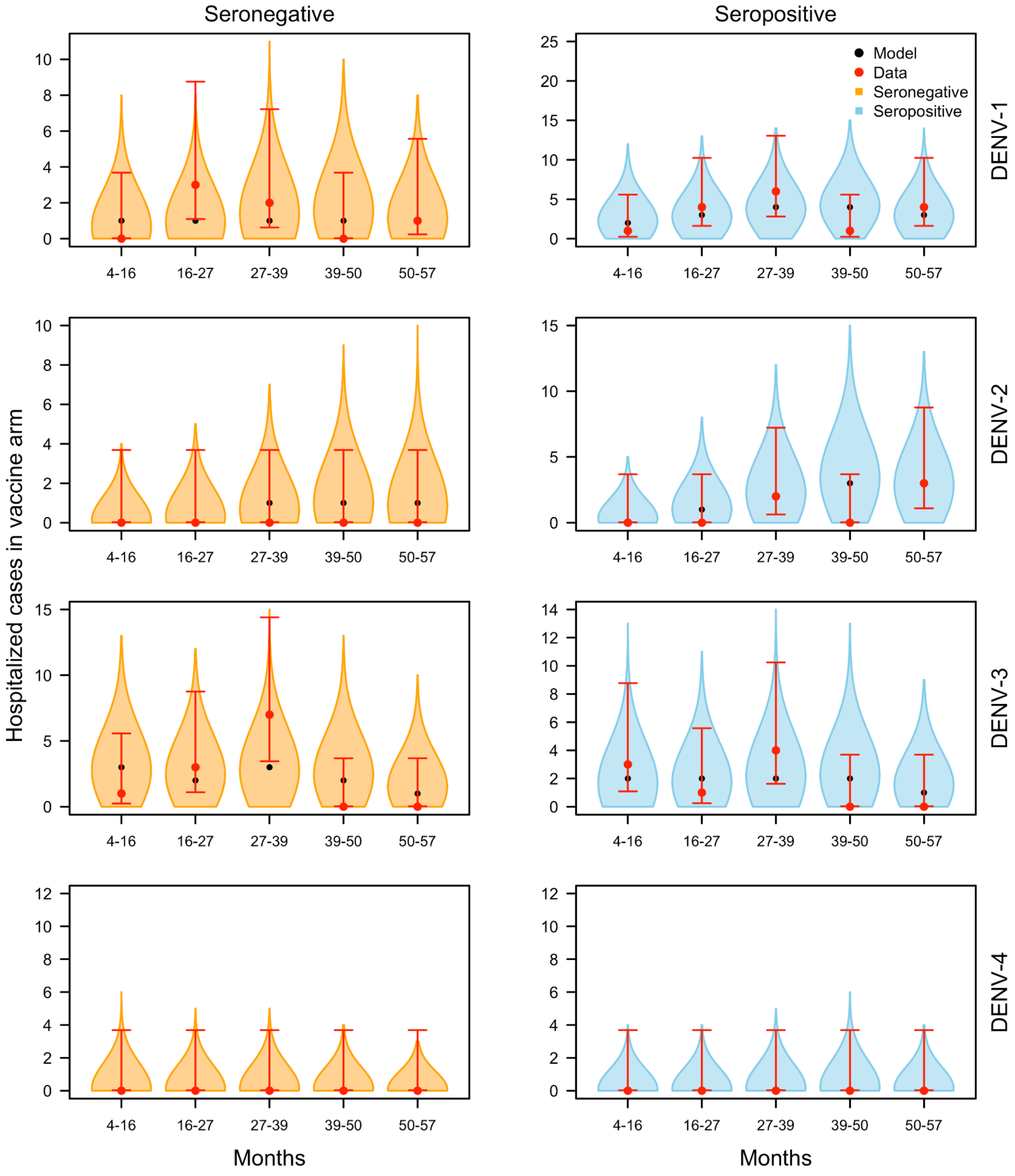


***Figure S15.*** *Posterior predictions of the number of hospitalized cases by serotype (rows) and serostatus (columns) from 4 to 57 months (x-axis) after the first vaccine dose in the vaccine arm over time. Violin shapes show how the model's posterior predictions were distributed, with black dots indicating median predictions. The red dot and lines show the observed value in the trial and the 95% posterior predictive interval around that observation, which was calculated assuming a beta conjugate prior (α=1, β=1) and a binomial likelihood to account for the sampling process and associated uncertainty involved in collecting the data.*

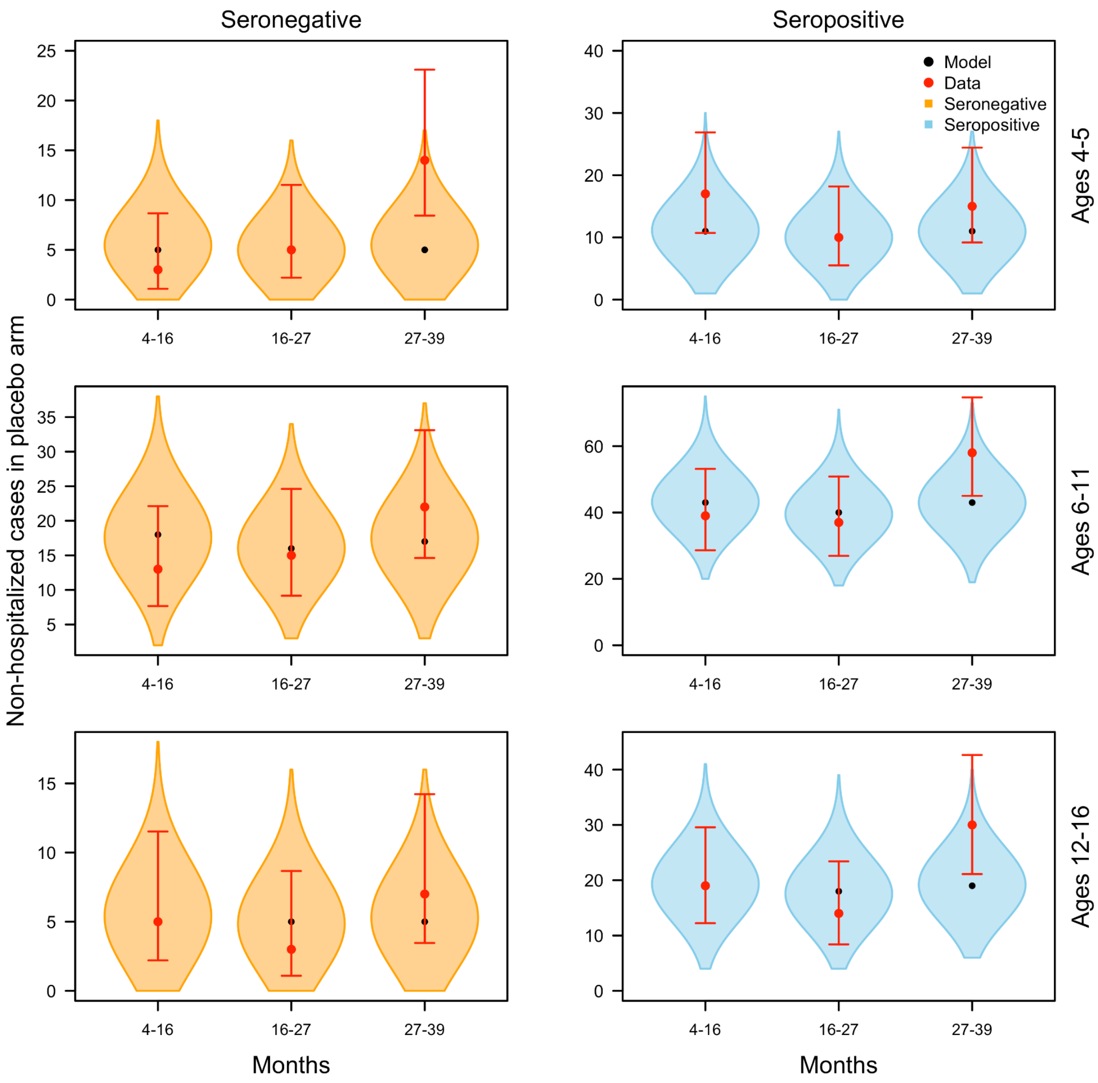


***Figure S16.*** *Posterior predictions of the number of non-hospitalized cases in each age group in the trial (rows) by serostatus (columns) from 4 to 57 months (x-axis) after the first vaccine dose in the placebo arm over time. Violin shapes show how the model's posterior predictions were distributed, with black dots indicating median predictions. The red dot and lines show the observed value in the trial and the 95% posterior predictive interval around that observation, which was calculated assuming a beta conjugate prior (α=1, β=1) and a binomial likelihood to account for the sampling process and associated uncertainty involved in collecting the data.*

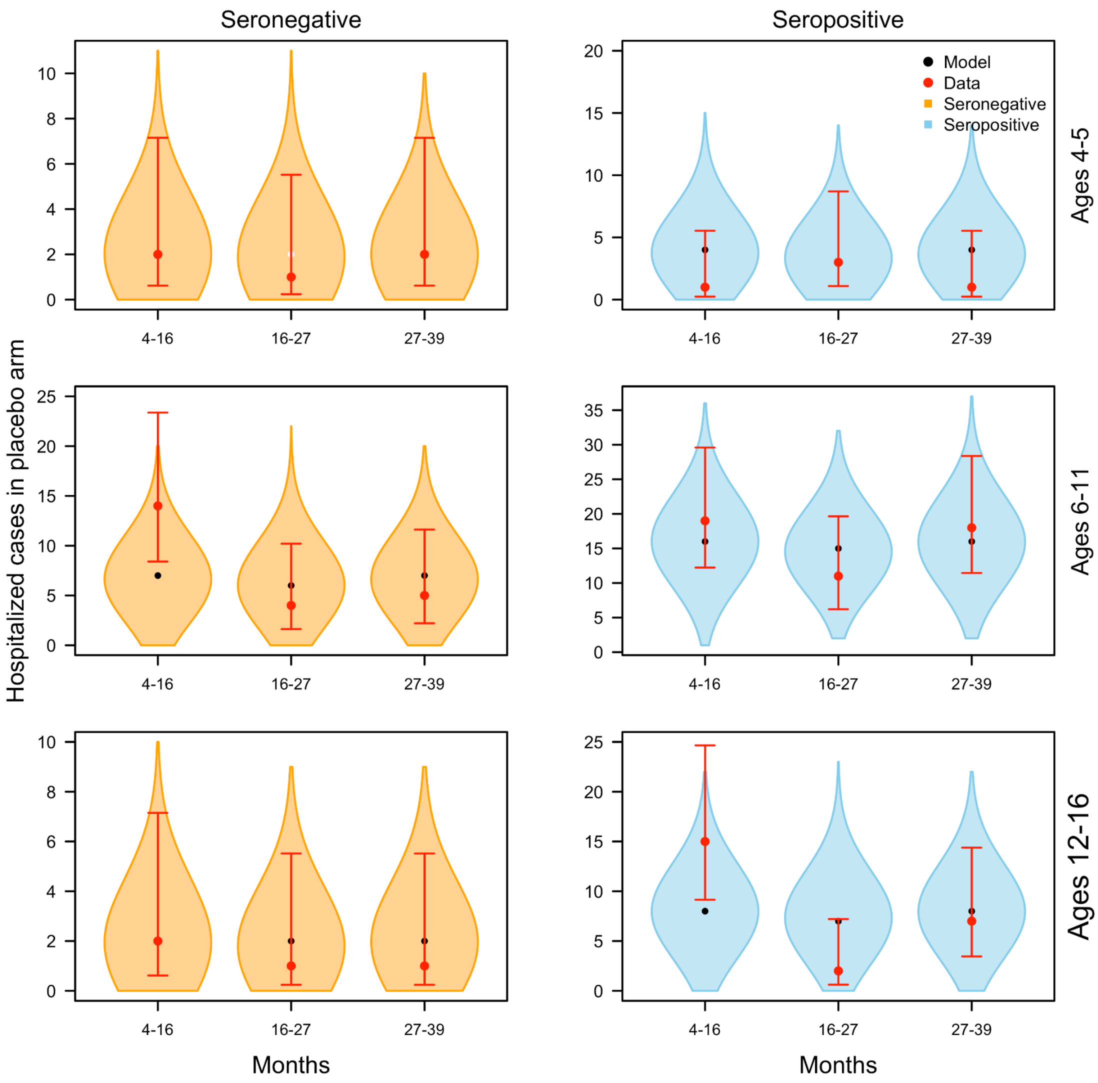


***Figure S17.*** *Posterior predictions of the number of hospitalized cases in each age group in the trial (rows) by serostatus (columns) from 4 to 57 months (x-axis) after the first vaccine dose in the placebo arm over time. Violin shapes show how the model's posterior predictions were distributed, with black dots indicating median predictions. The red dot and lines show the observed value in the trial and the 95% posterior predictive interval around that observation, which was calculated assuming a beta conjugate prior (α=1, β=1) and a binomial likelihood to account for the sampling process and associated uncertainty involved in collecting the data.*

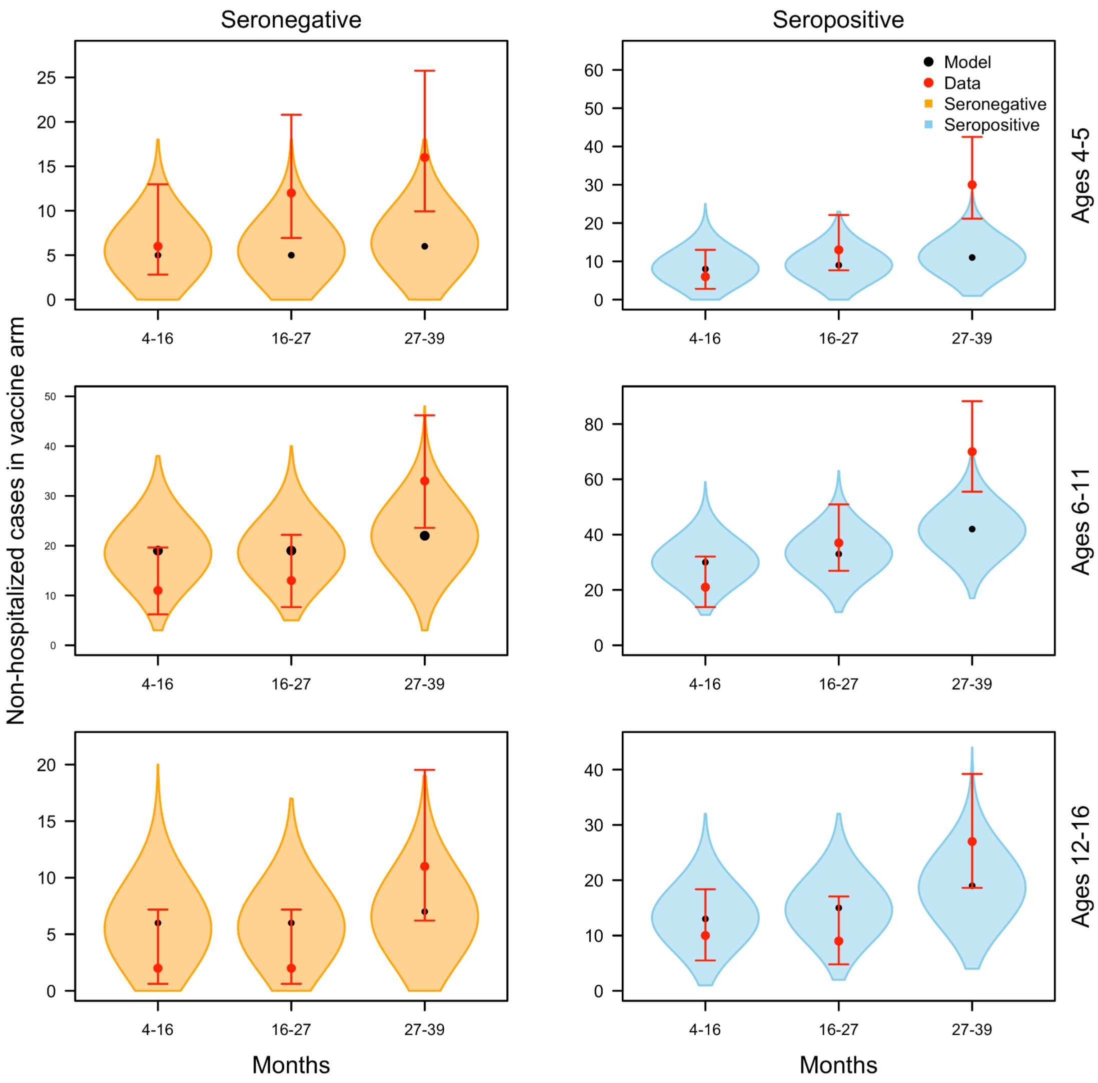


***Figure S18.*** *Posterior predictions of the number of non-hospitalized cases in each age group in the trial (rows) by serostatus (columns) from 4 to 57 months (x-axis) after the first vaccine dose in the vaccine arm over time. Violin shapes show how the model's posterior predictions were distributed, with black dots indicating median predictions. The red dot and lines show the observed value in the trial and the 95% posterior predictive interval around that observation, which was calculated assuming a beta conjugate prior (α=1, β=1) and a binomial likelihood to account for the sampling process and associated uncertainty involved in collecting the data.*

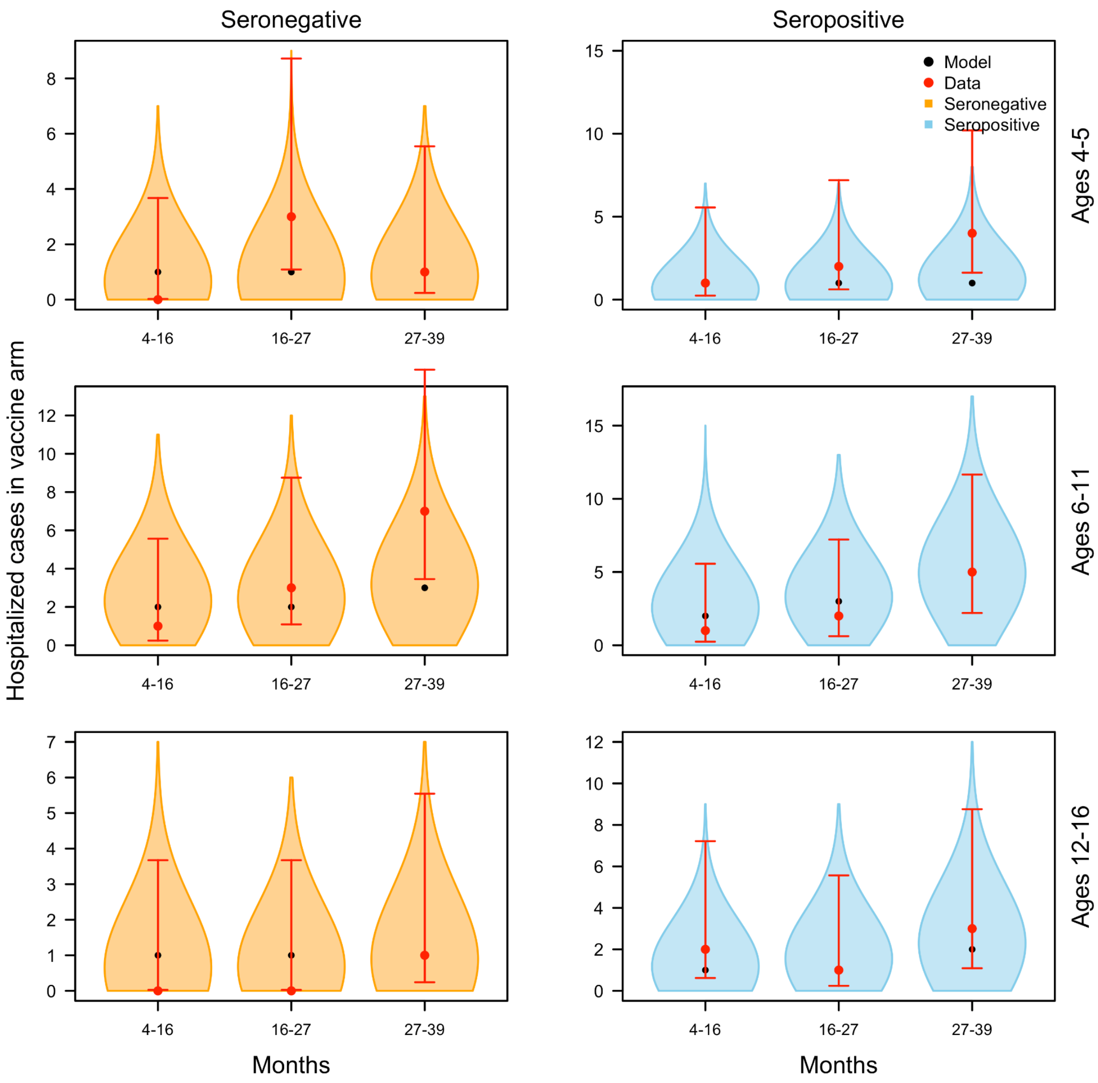


***Figure S19.*** *Posterior predictions of the number of hospitalized cases in each age group in the trial (rows) by serostatus (columns) from 4 to 57 months (x-axis) after the first vaccine dose in the vaccine arm over time. Violin shapes show how the model's posterior predictions were distributed, with black dots indicating median predictions. The red dot and lines show the observed value in the trial and the 95% posterior predictive interval around that observation, which was calculated assuming a beta conjugate prior (α=1, β=1) and a binomial likelihood to account for the sampling process and associated uncertainty involved in collecting the data.*

### 2.5 Model validation results

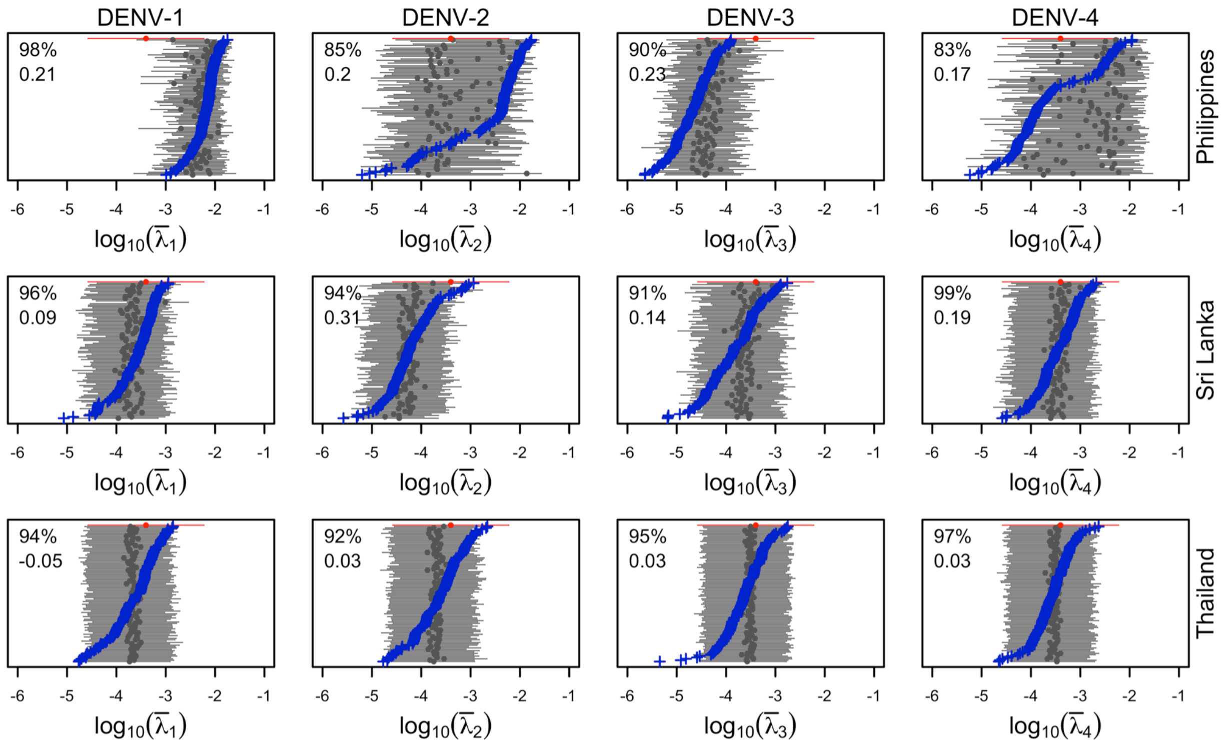


***Figure S20.*** *True parameter values (blue), recovered parameters and 95% credible intervals (gray), and prior distribution (red) for serotype-specific (columns) historical forces of infection for countries in Asia included in the trial (rows). In the upper left of each panel, the top value is coverage and the bottom value is the concordance correlation coefficient (see Methods of main text).*

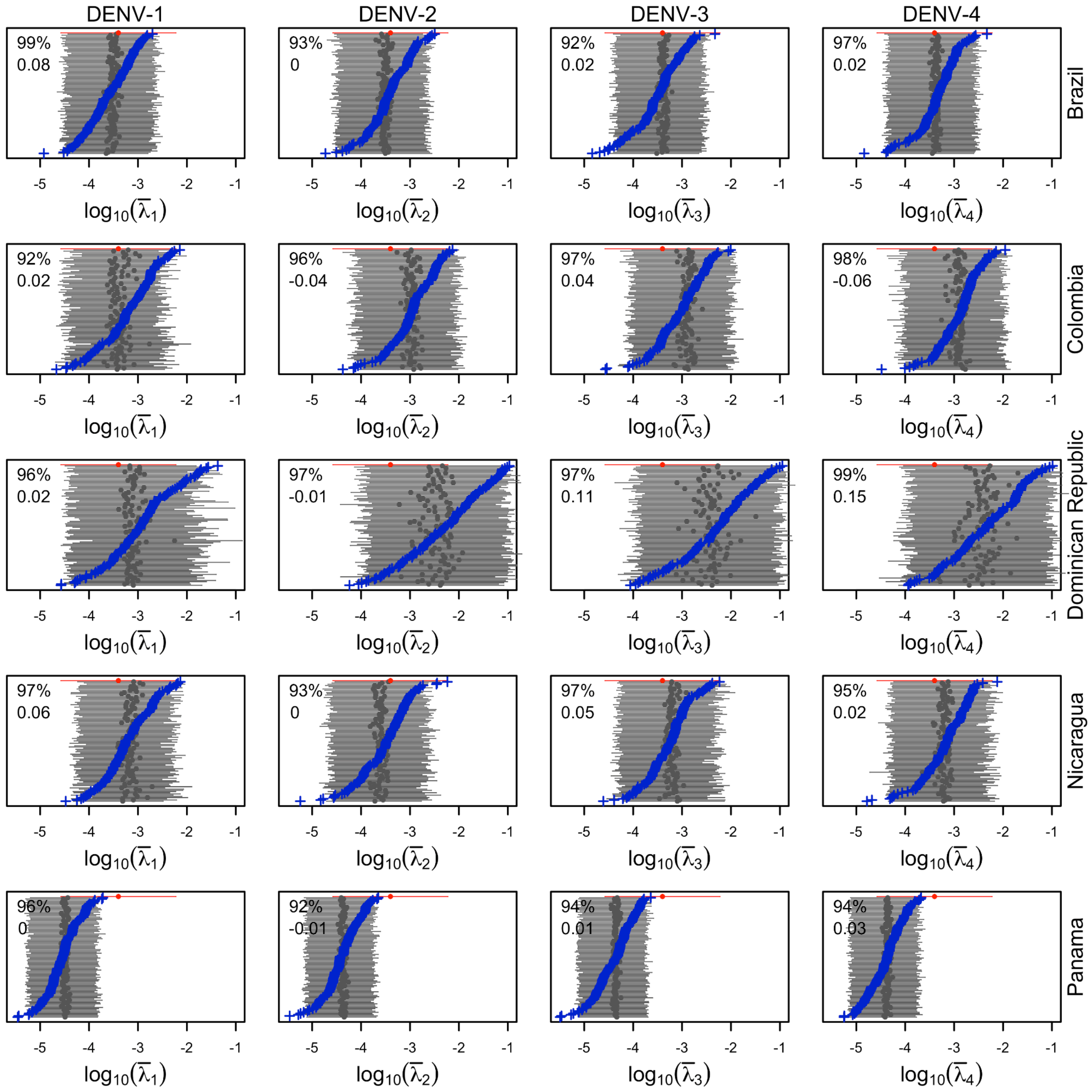


***Figure S21.*** *True parameter values (blue), recovered parameters and 95% credible intervals (gray), and prior distribution (red) for serotype-specific (columns) historical forces of infection for countries in Latin America included in the trial (rows). In the upper left of each panel, the top value is coverage and the bottom value is the concordance correlation coefficient (see Methods of main text).*

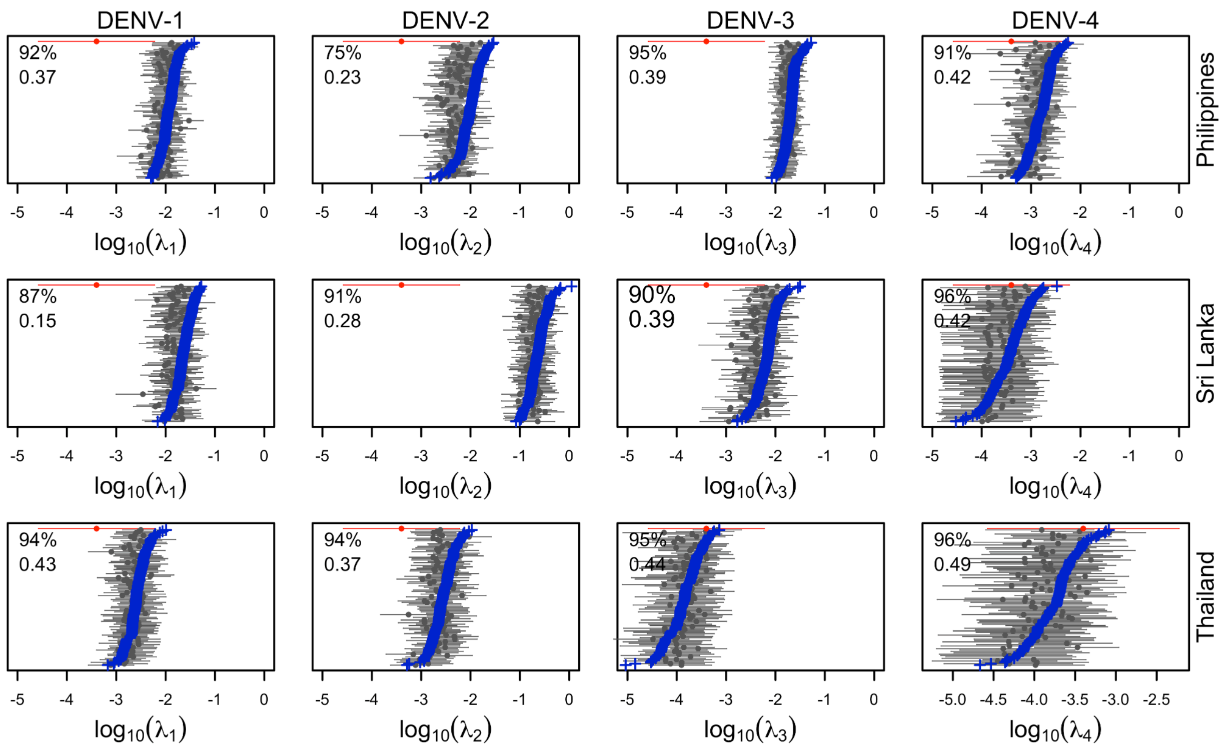


***Figure S22.*** *True parameter values (blue), recovered parameters and 95% credible intervals (gray), and prior distribution (red) for serotype-specific (columns) trial forces of infection for countries in Asia included in the trial (rows). In the upper left of each panel, the top value is coverage and the bottom value is the concordance correlation coefficient (see Methods of main text).*

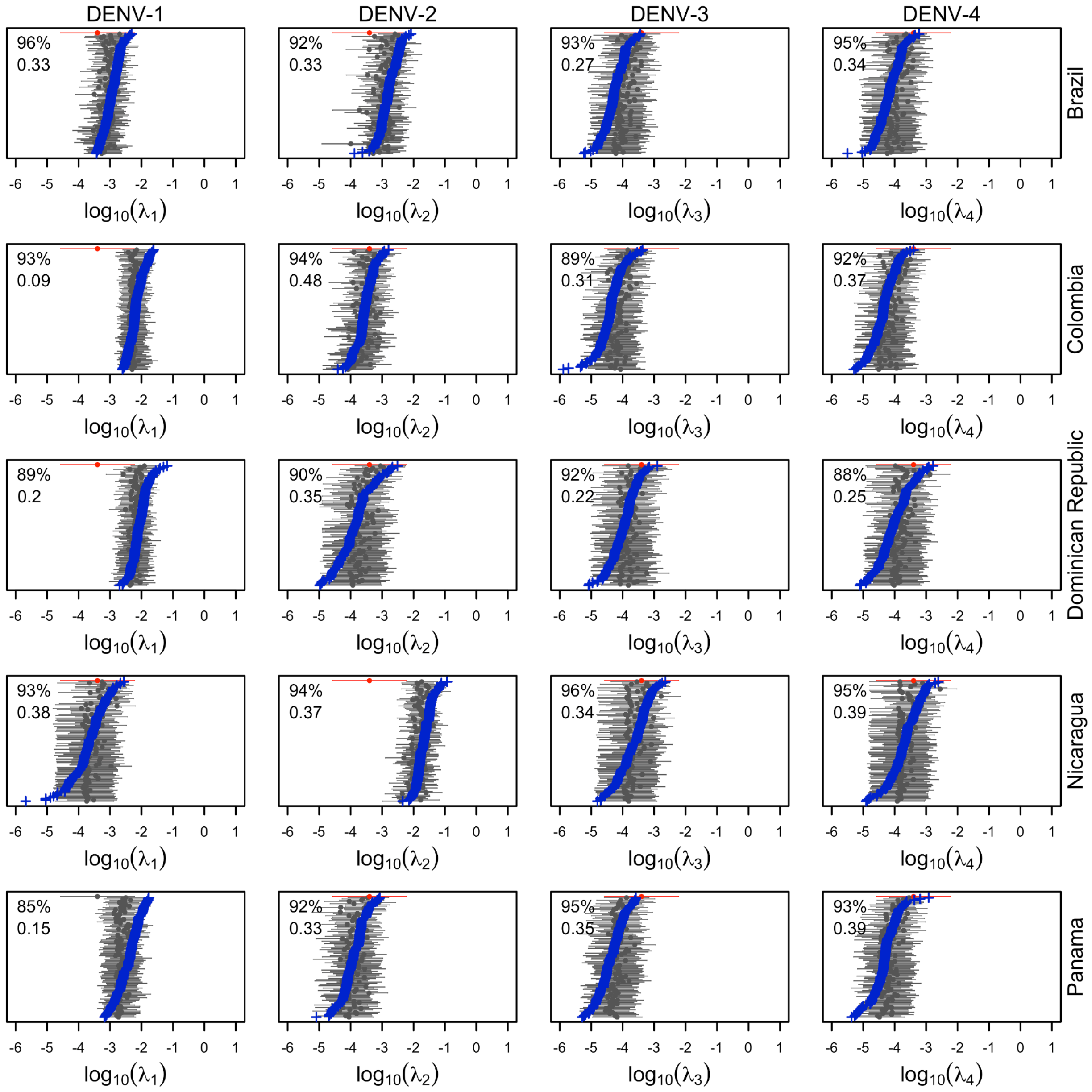


***Figure S23.*** *True parameter values (blue), recovered parameters and 95% credible intervals (gray), and prior distribution (red) for serotype-specific (columns) trial forces of infection for countries in Latin America included in the trial (rows). In the upper left, the top value is coverage and the bottom value is the concordance correlation coefficient (see Methods of main text).*

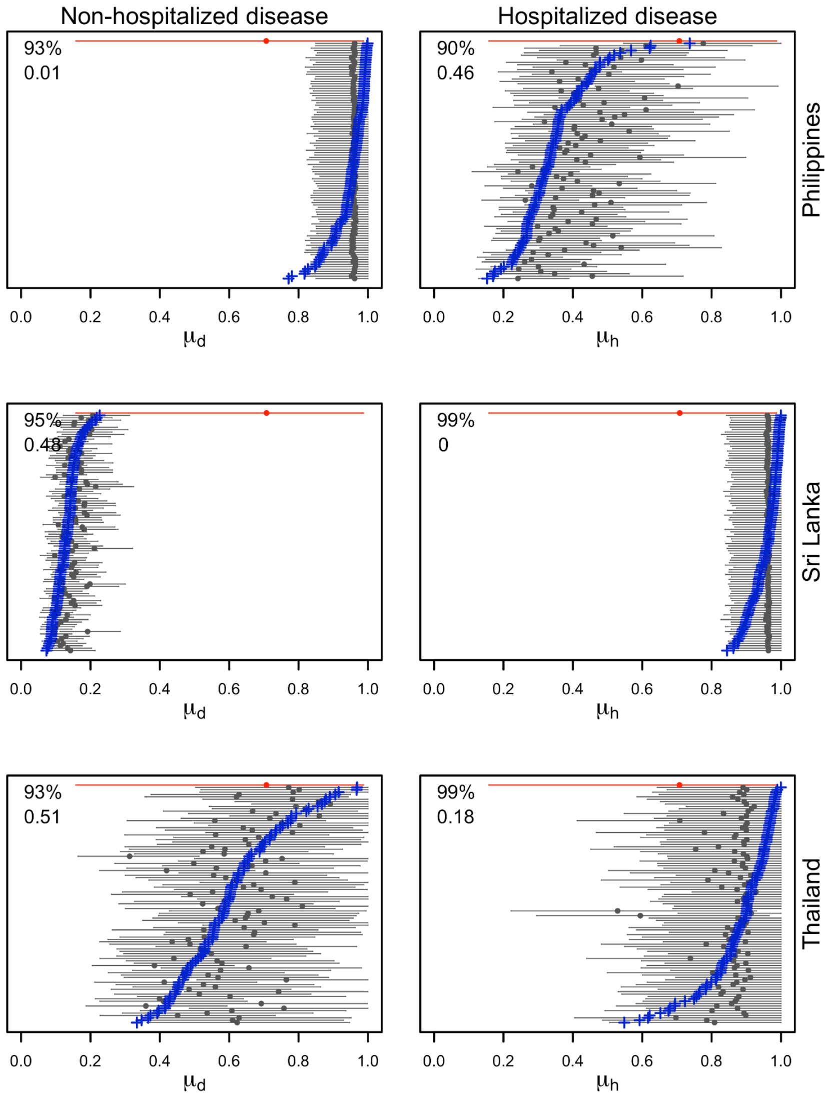


***Figure S24.*** *True parameter values (blue), recovered parameters and 95% credible intervals (gray), and prior distribution (red) for outcome-specific (columns) reporting probabilities for countries in Asia included in the trial (rows). In the upper left of each panel, the top value is coverage and the bottom value is the concordance correlation coefficient (see Methods of main text).*

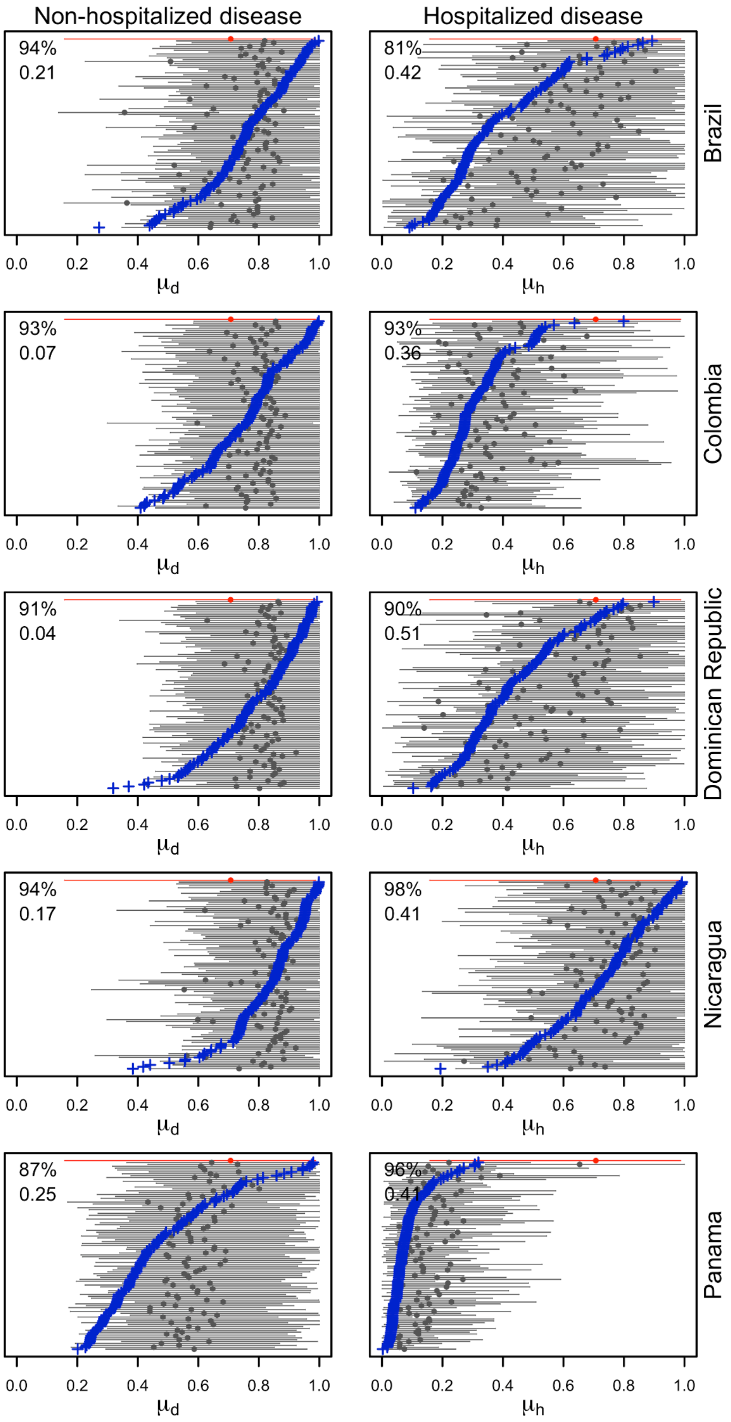


***Figure S25.*** *True parameter values (blue), recovered parameters and 95% credible intervals (gray), and prior distribution (red) for outcome-specific (columns) reporting probabilities for countries in Latin America included in the trial (rows). In the upper left of each panel, the top value is coverage and the bottom value is the concordance correlation coefficient (see Methods of main text).*

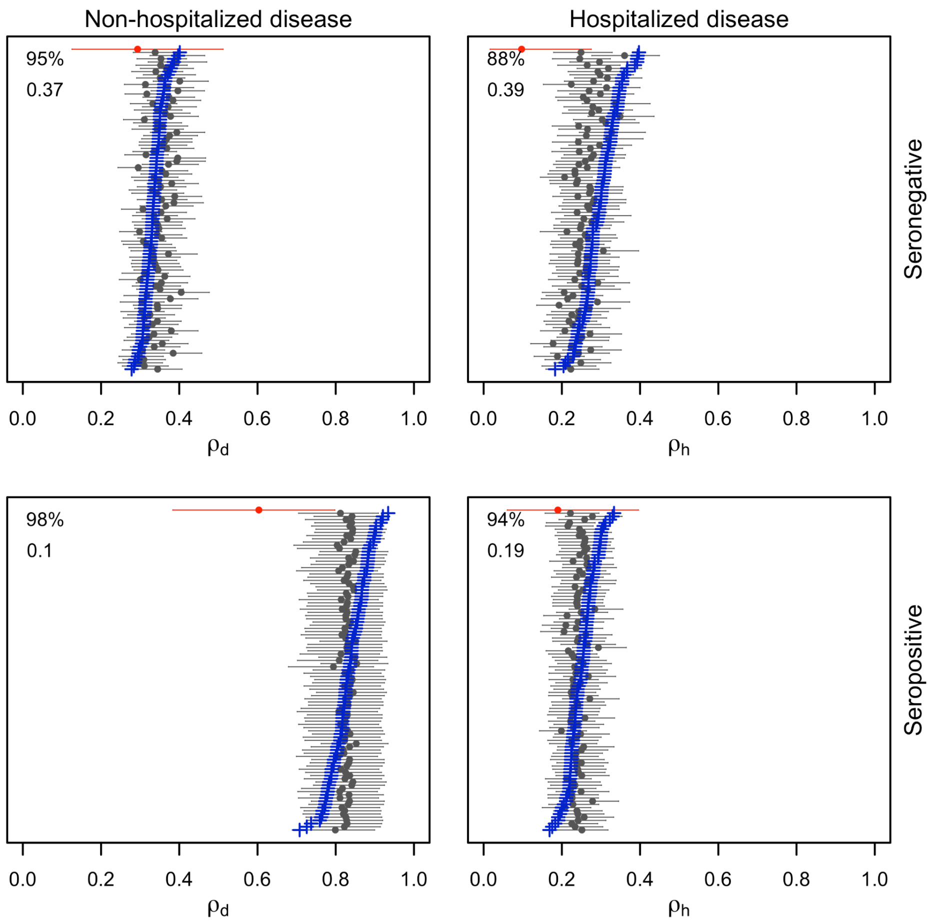


***Figure S26.*** *True parameter values (blue), recovered parameters and 95% credible intervals (gray), and prior distribution (red) for serostatus-specific (columns) outcome probabilities. In the upper left of each panel, the top value is coverage and the bottom value is the concordance correlation coefficient (see Methods of main text).*

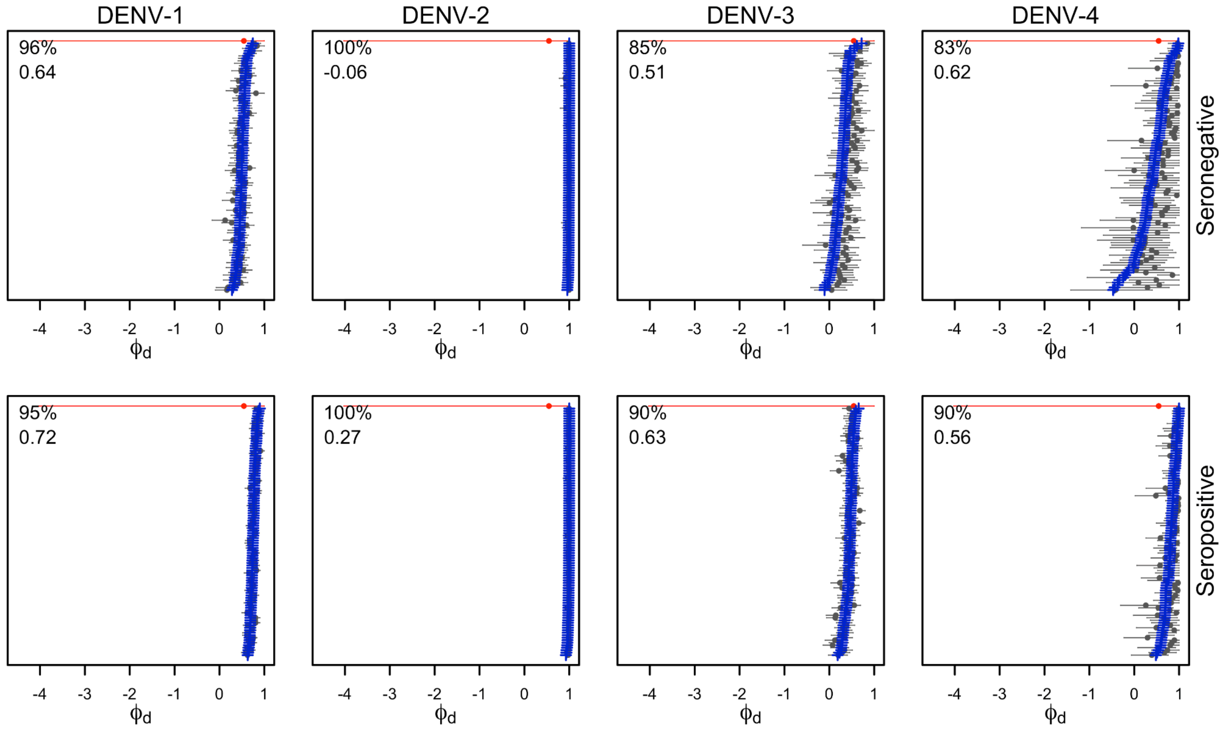


***Figure S27.*** *True parameter values (blue), recovered parameters and 95% credible intervals (gray), and prior distribution (red) for serostatus- (rows) and serotype-specific (columns) per-exposure protection against non-hospitalized disease. In the upper left of each panel, the top value is coverage and the bottom value is the concordance correlation coefficient (see Methods of main text).*

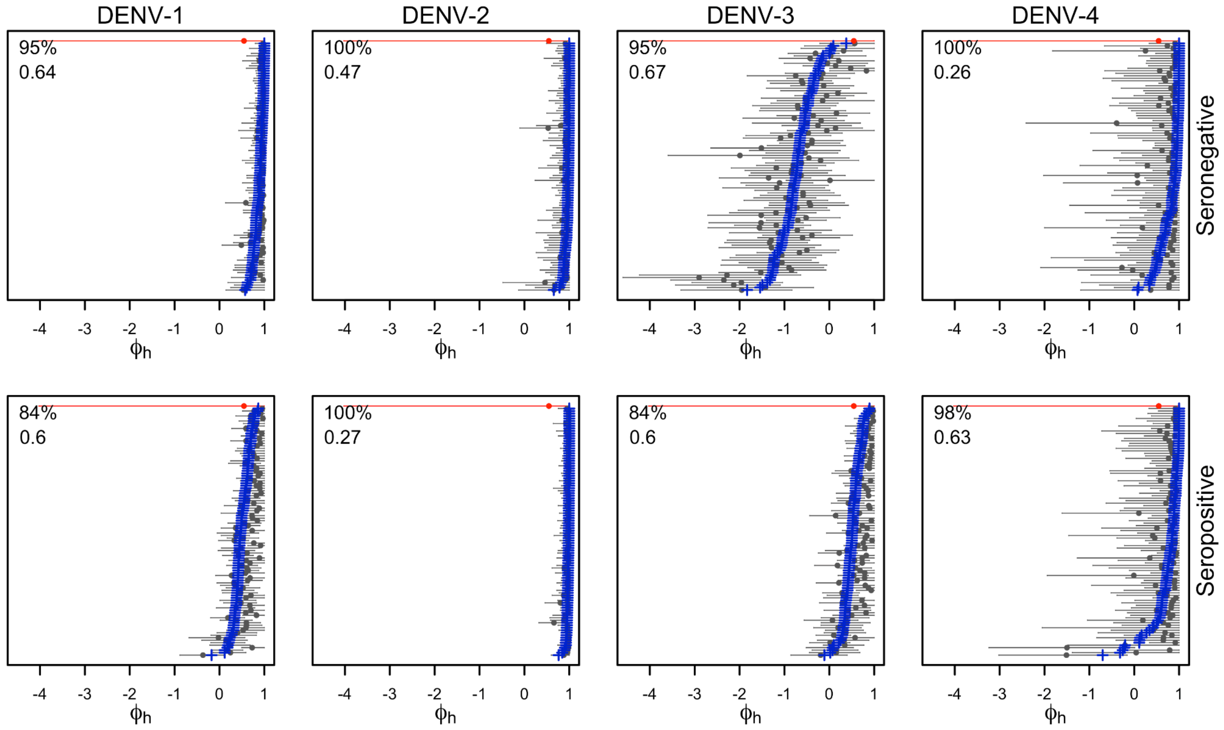


***Figure S28.*** *True parameter values (blue), recovered parameters and 95% credible intervals (gray), and prior distribution (red) for serostatus- (rows) and serotype-specific (columns) per-exposure protection against hospitalized disease. In the upper left of each panel, the top value is coverage and the bottom value is the concordance correlation coefficient (see Methods of main text).*

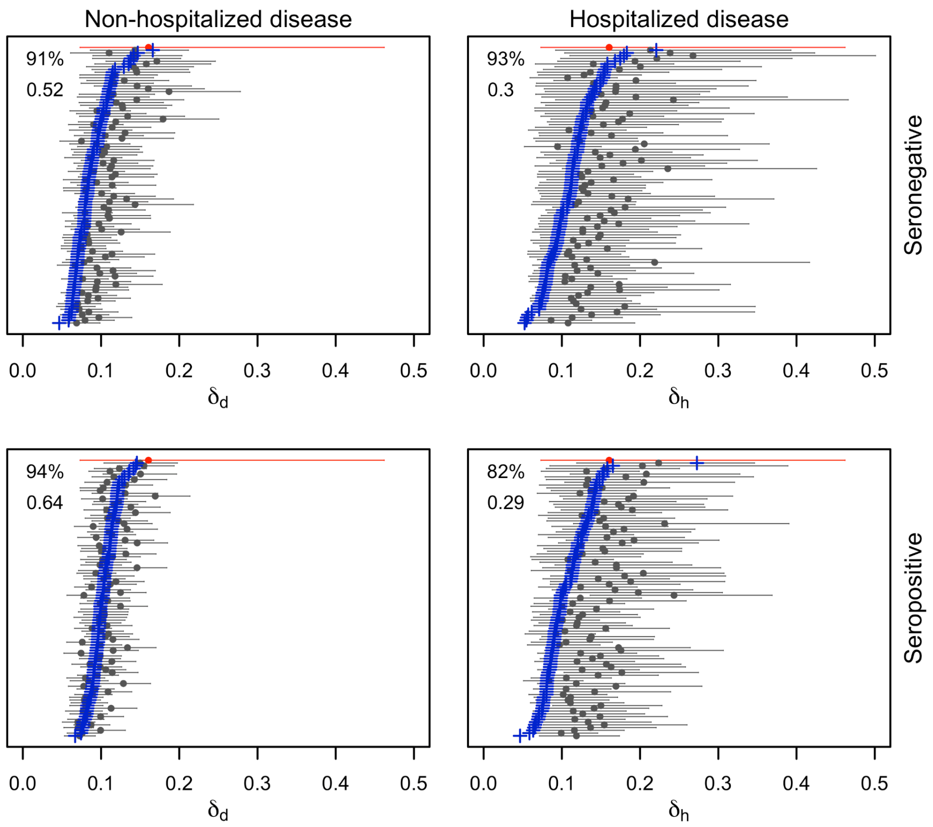


***Figure S29.*** *True parameter values (blue), recovered parameters and 95% credible intervals (gray), and prior distribution (red) for serostatus- (rows) and outcome-specific (columns) rates of waning of per-exposure protection. In the upper left of each panel, the top value is coverage and the bottom value is the concordance correlation coefficient (see Methods of main text).*
